# Association between SARS-CoV-2 Transmissibility, Viral Load, and Age in Households

**DOI:** 10.1101/2021.02.28.21252608

**Authors:** Frederik Plesner Lyngse, Kåre Mølbak, Kristina Træholt Franck, Claus Nielsen, Robert Leo Skov, Marianne Voldstedlund, Arieh S. Cohen, Carsten Thure Kirkeby

## Abstract

**Aim:** The objective of this nationwide study was to investigate the association between SARS-CoV-2 transmissibility, viral load, and age of primary cases in Danish households.

**Background:** Spread in households represents a major mode of transmission of SARS-CoV-2. In order to take proper action against the spread of the disease, it is important to have a better understanding of transmission in the household domain—including the role of viral load of primary cases.

**Methods:** The study was designed as an observational cohort study, using detailed administrative register data. We included the full population of Denmark and all SARS-CoV-2 tests (August 25, 2020 to February 10, 2021) to estimate transmissibility in households comprising 2-6 people. RT-PCR Cycle threshold (Ct) values were used as a proxy for viral load.

**Results:** We identified 63,657 primary cases and 139,882 household members of which 21% tested positive by RT-PCR within a 1-14 day period after the primary case. There was an approximately linear association between Ct value of the sample and transmissibility, implying that cases with samples having a higher viral load were more transmissible than cases with samples having a lower viral load. However, even for primary cases with relatively high sample Ct values, the transmissibility was not negligible, e.g., for primary cases with a sample Ct value of 38, we found that 13% of the primary cases had at least one secondary household case. Moreover, 34% of all secondary cases were found in households with primary cases having sample Ct values >30. An increasing transmissibility with age of the primary cases for adults (≥20 years) and a decreasing transmissibility with age for children (<20 years) were found.

**Conclusions:** Although primary cases with sample high viral loads (low Ct values) were associated with higher SARS-CoV-2 transmissibility, we found no obvious cut-off for sample Ct values to eliminate transmissibility and a substantial amount of household transmission occurred in households where the primary cases had high sample Ct values (low viral load), The study further showed that transmissibility increases with age. These results have important public health implications, as they suggest that contact tracing should prioritize cases according to Ct values and age, and underline the importance of quick identification and isolation of cases. Furthermore, the study highlights that households can serve as a transmission bridge by creating connections between otherwise separate domains.

## 1 Introduction

The world is in the midst of the pandemic caused by the Severe Acute Respiratory Syndrome Coronavirus 2 (SARS-CoV-2). As the pandemic has huge medical as well as economic consequences, it is essential to understand the SARS-CoV-2 transmission dynamics and associated factors in order to improve interventions to control the spread of the disease, such as contact tracing efforts. Close person-to-person contact represents a main mode of transmission and households are one of the most important domains where such close contacts occur. Elucidation of risk factors for transmission within households is therefore of major importance.

Real-time reverse transcription polymerase chain reaction (RT-PCR) from nasopharyngial and oropharyngial swabs is used worldwide to detect SARS-CoV-2 (Corman et al., 2020). Detection of SARS-CoV-2 by RT-PCR is—similar to other viruses—dependent on the viral load and is reversely correlated with the Cycle threshold (Ct) value of the test (Singanayagam et al., 2020). The viral load changes during the course of disease and has been shown to be highest around the time of symptom onset (He et al., 2020).

Recently, studies have shown that higher viral load (low Ct values) was associated with increased transmissibility of SARS-CoV-2, and it has been argued that contact tracing should be prioritized on cases with a high viral load, as they will have a higher risk of generating secondary cases (Marks et al., 2021; Lee et al., 2021a,b). Furthermore, it has also been described that transmissibility depends on the age of the infected individual and the age of the exposed individual (Lyngse et al., 2020, 2021; Lee et al., 2021a,b).

The aim of this nationwide observational study was to investigate the association between the transmissibility of SARS-CoV-2, RT-PCR Ct values, while taking age of infected cases into account.

## 2 Data and Methods

In Denmark all residents have access to tax-paid universal health insurance and tests for SARS-CoV-2 are free of charge. The testing capacity has increased during the pandemic and is widespread. Furthermore, SARS-CoV-2 sick leave is fully reimbursed by the state. Thus, neither access to tests nor financial reasons were major obstacles to obtaining a test during our study period. RT-PCR tests for SARS-CoV-2 could be obtained from either community testing facilities at TestCenter Denmark (TCDK) or from clinical microbiology laboratories at hospitals. Statens Serum Institut (SSI) analyses all tests from TCDK. Information on Ct values was only available for samples that tested positive at TCDK.

### 2.1 Register Data

In this study, we used Danish administrative register data comprising the full population. All residents in Denmark have a unique personal identification number that allows a complete linkage of information across different registers at the individual level. All results from RT-PCR tests for SARS-CoV-2 in Denmark are registered in the Danish Microbiology Database (MiBA), from which we obtained data on the individual level for all national tests for SARS-CoV-2 during the period August 25, 2020 (which was the first date with accessible Ct values) to February 10, 2021. Primary cases were only included until January 25, 2021 in order to allow for secondary cases to present within the following 14 days (see definition of cases below). Moreover, we only included primary cases identified by TCDK, as we only had Ct values on those case samples. To identify secondary cases, we included all tests, regardless of whether they tested at TCDK or hospitals.

Information on the reason for being tested (e.g., symptoms, potential contact with infected persons etc.) was not available. From the Danish Civil Registry System, we obtained information about the sex, age, and home address for all individuals living in Denmark. People who tested positive for SARS-CoV-2 using antigen tests were not included, as these test results were not transferred to MiBA at the time of the study.

### 2.2 Data Linkage

We constructed households by linking all individuals living at the same address, including identification of apartments (e.g., six single apartments in the same building counted as six independent households). Only households with two to six members were included in the study in order to exclude, e.g., residential institutions, such as long term care facilities. Person-level data, including information on the test result, the date of sampling, and the date the result was available, was linked to individuals within each household. For every household, we identified the person with the first positive test for SARS-CoV-2; referred to as the primary case throughout this paper. If there was more than one possible primary case, i.e., if two or more cases tested positive on the same day—and were the first identified cases in the household—one was randomly assigned as the primary case. We considered all subsequent tests from other members in the same household as being tests taken in response to the primary case and defined secondary cases as those who had a positive test with sampling dates 1 to 14 days after the primary case tested positive. In addition, we assumed that all identified secondary cases were infected by the primary case within the same household. Lastly, we refer to the Ct value of the first positive sample for each primary case (i.e., the identifying sample) as Ct value throughout the paper.

### 2.3 Laboratory Analyses

Analysis of tests at TCDK was performed using a set of primers that target the E-gene on SARS-CoV-2 (Corman et al., 2020), which is recommended by the WHO and the ECDC, and has a high sensitivity and specificity (Vogels et al., 2020). TCDK have used the same methodology and primers throughout the epidemic, making the Ct values comparable across the study period. An RT-PCR test for SARS-CoV-2 was defined as positive if the Ct value was ≤38 (SSI, 2021).

### 2.4 Statistical Analyses

We utilized two concepts for transmissibility of the primary case: transmission risk and transmission rate, as described in Lyngse et al. (2021). The *transmission risk* describes the risk of infecting at least one other person within the household, and equals one if any (one or more) secondary cases are identified within the same household, and zero otherwise. The *transmission rate* is the proportion of potential secondary cases within the same household that tested positive. Both transmissibility measures were weighted on the primary case level, such that each primary case has a weight of one.

In addition, we utilized one concept for the susceptibility of the potential secondary case: attack rate. The (secondary) attack rate is defined as the proportion of potential secondary cases that tested positive. The attack rate was weighted on the potential secondary case level such that each potential secondary case has a weight of one.

We estimated the association between Ct values and transmissibility for each Ct value separately, using a non-parametric linear regression.

We grouped primary cases according to their Ct values, and we counted their associated positive secondary cases. From this, we calculated the proportion of secondary cases that were associated with the identification of primary cases with a Ct value above a certain threshold.

We estimated the association between age and transmissibility stratified by the median Ct value for each five-year age group separately using a non-parametric linear regression.

We estimated the association between Ct values, age, and transmissibility. We grouped primary cases according to their Ct values in bins of two and ten-year age groups, and we estimated the transmissibility separately using a non-parametric linear regression.

We used a logistic regression model to estimate the odds ratio of the transmissibility for Ct value, age, sex, and household size. We used a univariable model to investigate the effect of each explanatory variable separately, and a multivariable model to investigate the combined effect.

#### 2.4.1 Sensitivity analyses

We estimated the age structured transmissibility stratified by sex to investigate whether there were different patterns across men and women. We also estimated the age structured transmissibility stratified by Ct value quartiles to investigate whether the pattern was independent of the dichotomization using the median Ct value. As Ct values were only available for the primary cases that were identified by being tested at TCDK, there is a potential bias from not including primary cases identified at hospitals. We, therefore, estimated the age structured transmissibility stratified by place of testing.

Because people normally live with a partner around their own age and parents live with their children, susceptibility correlation with age could drive an age structured transmissibility. To address this potential bias, we stratified our sample by Ct value quartiles and estimated the transmission rate with the interaction of the age of the primary and potential secondary case.

Lastly, one could think that primary cases with high Ct values (low viral load) were due to sampling late in the infection and cases classified as secondary cases were actually primary or co-primary cases. To address this concern, we investigated whether changing the criteria for including secondary cases in relation to time of identification of the primary case, i.e., only including secondary cases found positive on days 1-14 (as in the main analyses), 2-14, 3-14, and 4-14, respectively, affected the results.

### 2.5 Ethical statement

This study was conducted using administrative register data. According to Danish law, ethics approval is not needed for such research. All data management and analyses were carried out on the Danish Health Data Authority’s restricted research servers with project number FSEID-00004942.

## 3 Results

### 3.1 Descriptive Statistics

From August 25, 2020 to February 10, 2021, TCDK analyzed 9,347,218 samples for SARS-CoV-2 (75% of all tests in Denmark) and identified 63,937 primary cases up to January 25 (73% of all primary cases in Denmark) (Table S1). A Ct value was available for 99.6% of these primary cases.

A total of 63,657 primary cases with a Ct value were registered as living in households comprising of 2-6 people with 139,882 potential secondary cases, of which 29,739 tested positive for SARS-CoV-2 1-14 days after the primary case was tested and were thus included as positive secondary cases in this study. This implies an attack rate of 21% (29,739/139,882). Across primary cases, potential secondary cases and positive secondary cases, the distribution of sex, age, household size, attack rate, and Ct values of the primary cases are shown in Table 1. (See Table S2 for 5-year age distributions and Table S3 for the distribution of Ct values of primary cases, including attack rates.)

**Table 1:** Summary statistics for the study data

|  | Primary<br>Cases | Potential<br>Secondary<br>Cases | Positive<br>Secondary<br>Cases | Attack<br>Rate<br>(%) |
| --- | --- | --- | --- | --- |
| **Total** | 63,657 | 139,882 | 29,739 | 21 |
| **Sex** |  |  |  |  |
| Male | 32,256 | 70,849 | 14,646 | 21 |
| Female | 31,401 | 69,002 | 15,090 | 22 |
| **Age** |  |  |  |  |
| 0-10 | 3,711 | 25,015 | 4,733 | 19 |
| 10-20 | 14,280 | 29,277 | 5,847 | 20 |
| 20-30 | 13,432 | 20,137 | 3,705 | 18 |
| 30-40 | 8,814 | 15,034 | 3,601 | 24 |
| 40-50 | 9,393 | 24,854 | 5,120 | 21 |
| 50-60 | 8,781 | 17,795 | 4,150 | 23 |
| 60-70 | 3,650 | 5,375 | 1,719 | 32 |
| 70-80 | 1,423 | 2,021 | 737 | 36 |
| >80 | 173 | 343 | 124 | 36 |
| **Household Size** |  |  |  |  |
| 2 | 21,830 | 21,830 | 6,345 | 29 |
| 3 | 15,226 | 29,375 | 5,948 | 20 |
| 4 | 16,773 | 48,532 | 9,725 | 20 |
| 5 | 7,528 | 29,066 | 5,642 | 19 |
| 6 | 2,300 | 11,079 | 2,079 | 19 |
| **Ct Value** |  |  |  |  |
| <19 | 360 |  |  |  |
| 19-20 | 1,486 |  |  |  |
| 21-22 | 4,328 |  |  |  |
| 23-24 | 7,289 |  |  |  |
| 25-26 | 9,168 |  |  |  |
| 27-28 | 9,533 |  |  |  |
| 29-30 | 8,650 |  |  |  |
| 31-32 | 7,391 |  |  |  |
| 33-34 | 6,381 |  |  |  |
| 35-36 | 5,397 |  |  |  |
| 37-38 | 3,674 |  |  |  |

The distribution of Ct values in samples from primary cases is shown in Figure S1: 25% of primary cases had a Ct value ≤25, 50% had a Ct value ≤28, and 75% had a Ct value ≤32. The distribution of Ct values was relatively similar across age groups, suggesting that differences in test strategies across age were not driving the results of this study (Figure S2 and Table S4).

### 3.2 Associations with transmissibility

Figure 1 shows the association between Ct values and transmissibility. There was an approximately linear decreasing relationship between Ct values and transmissibility. Both the transmission rate (blue) and the transmission risk (red) decreased with an increasing Ct value of the primary case. A primary case with a Ct value of 18 had a transmission rate of 47% and a transmission risk of 58%, compared to a transmission rate of 9% and a transmission risk of 13% for a Ct value of 38.

**Figure 1:**
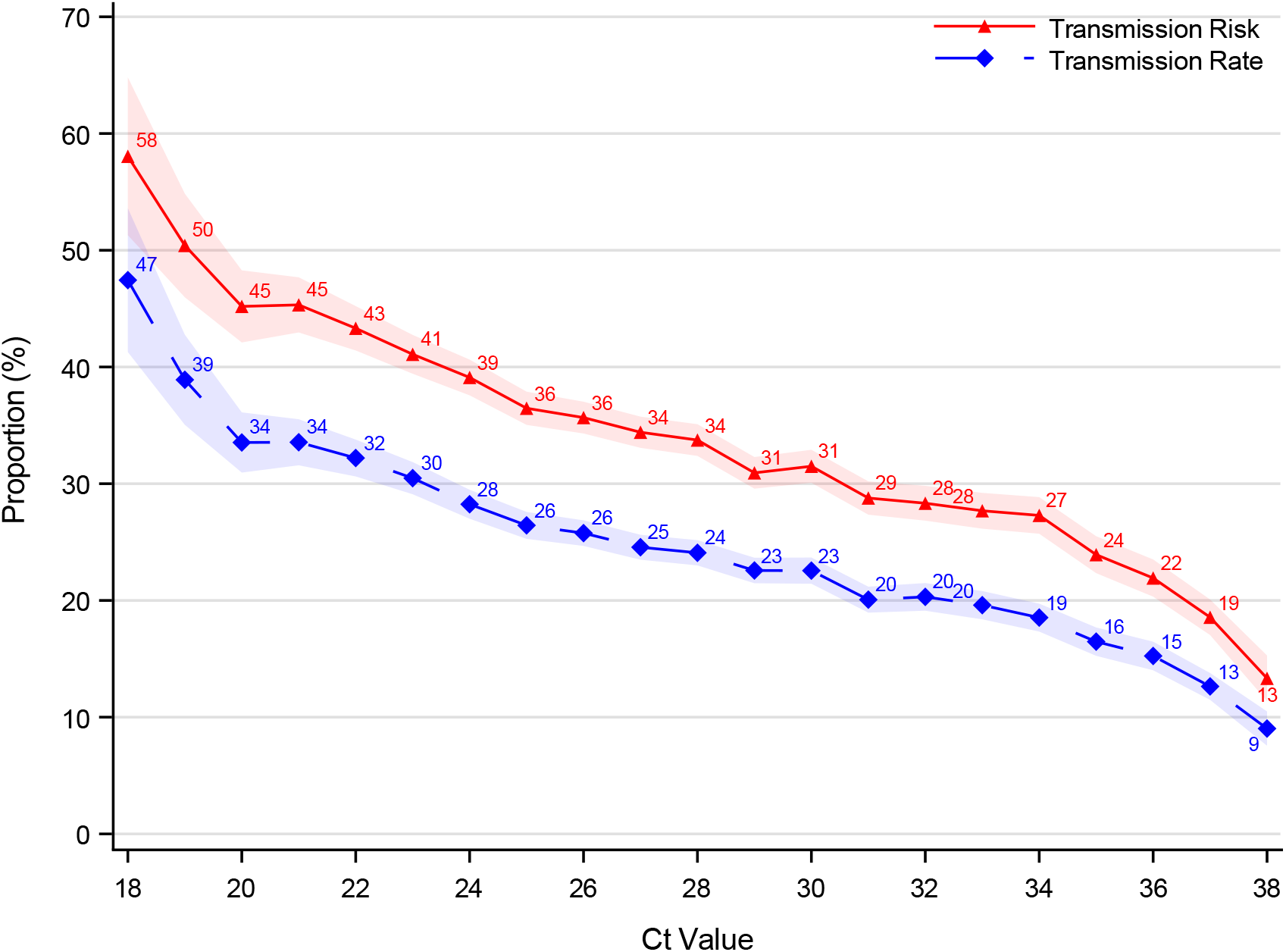
Association between Ct values and transmissibility Notes: The figure shows the association between sample Ct values and transmissibility. The transmission rate (blue) describes the proportion of potential secondary cases within the household that were infected. The transmission risk (red) describes the proportion of infected primary cases that infected at least one secondary case. A primary case with a sample Ct value of 18 had a transmission rate of 47% and a transmission risk of 58%, compared to a transmission rate of 9% and a transmission risk of 13% for a Ct value of 38. An RT-PCR test is positive if the Ct value is ≤38. The shaded area shows the 95% confidence bands clustered on the household level.

Figure 2 shows the reverse cumulative distribution of secondary cases by the Ct value of the primary cases. E.g., primary cases with a Ct value ≥30 account for 34% of the total positive secondary cases within this study.

**Figure 2:**
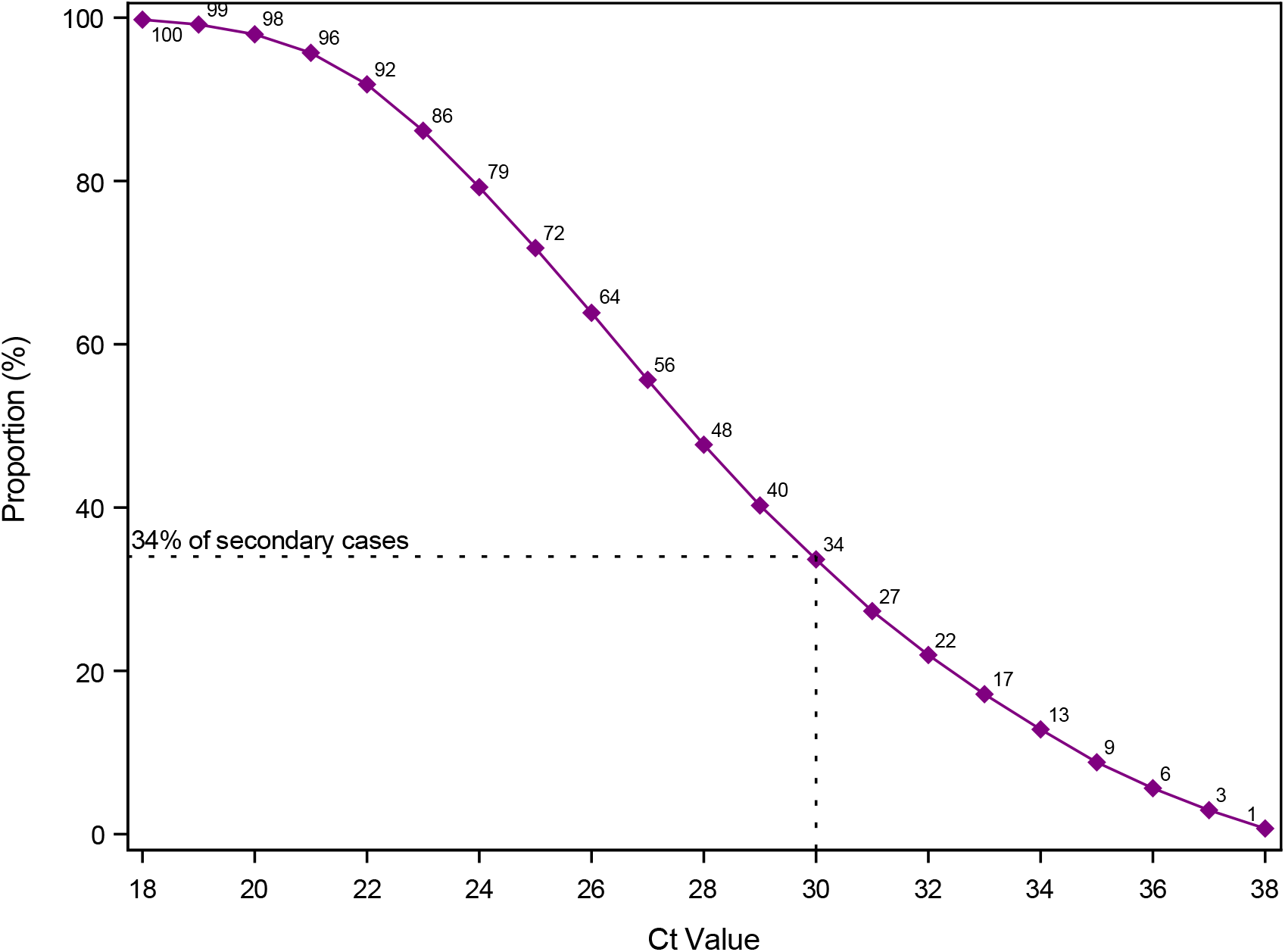
Proportion of secondary cases associated with Ct values of primary cases above a threshold Notes: The figure shows the reverse cumulative distribution of secondary cases by sample Ct value of the primary cases. E.g., primary cases with a sample Ct value ≥30 account for 34% of the total positive secondary cases within this study. An RT-PCR test is positive if the Ct value is ≤38.

Figure 3 shows the association between age and transmissibility stratified by the median Ct value (Ct=28). Primary cases with a Ct value below the median (red) were significantly more transmissible than primary cases with a Ct value above the median (blue), across all age groups.

**Figure 3:**
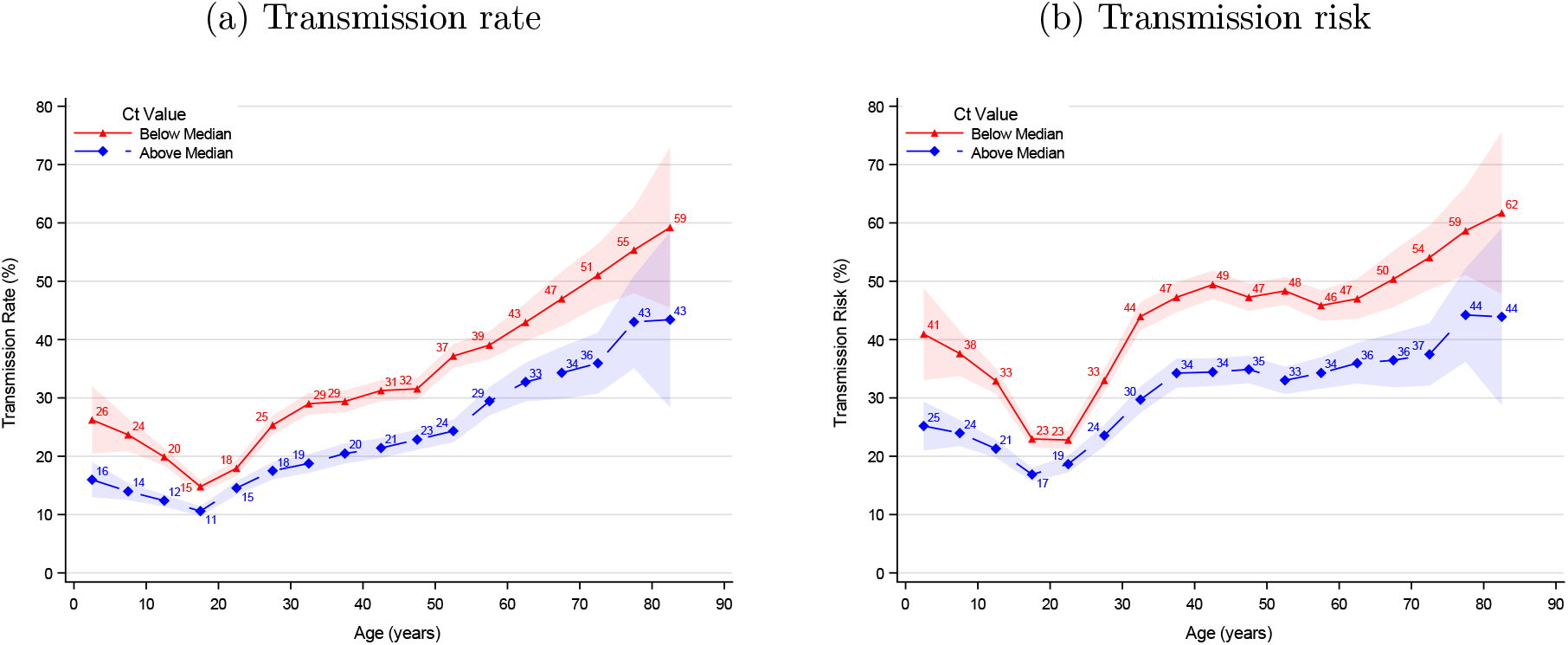
Age structured transmissibility stratified by median Ct value Notes: This figure shows the association between age and transmissibility stratified by median sample Ct value (Ct=28). The transmission rate describes the proportion of potential secondary cases within the household that were infected. The transmission risk describes the proportion of infected primary cases that infected at least one secondary case. A primary case aged 0-5 years with a sample Ct value <28 had a transmission rate of 26%, while a primary case aged 0-5 years with a sample Ct value ≥28 had a transmission rate of 16% (panel a). For the transmission risk, these estimates were 41% and 25%, respectively (panel b). The shaded areas show the 95% confidence bands clustered on the household level.

Primary cases aged 15-20 years had the lowest transmissibility. Younger children had a higher transmissibility compared to older children, whereas older adults had a higher transmissibility compared to younger adults.

Figure 4 shows the association between age of the primary case, Ct value and transmissibility. Within each age group the transmissibility increased with decreasing Ct values. Within Ct value intervals, the transmission rate generally increased with the age of the primary case, except for children aged 0-10 years. Adults aged 30+ and children aged 0-10 years generally had a higher transmission risk compared to persons aged 10-30.

**Figure 4:**
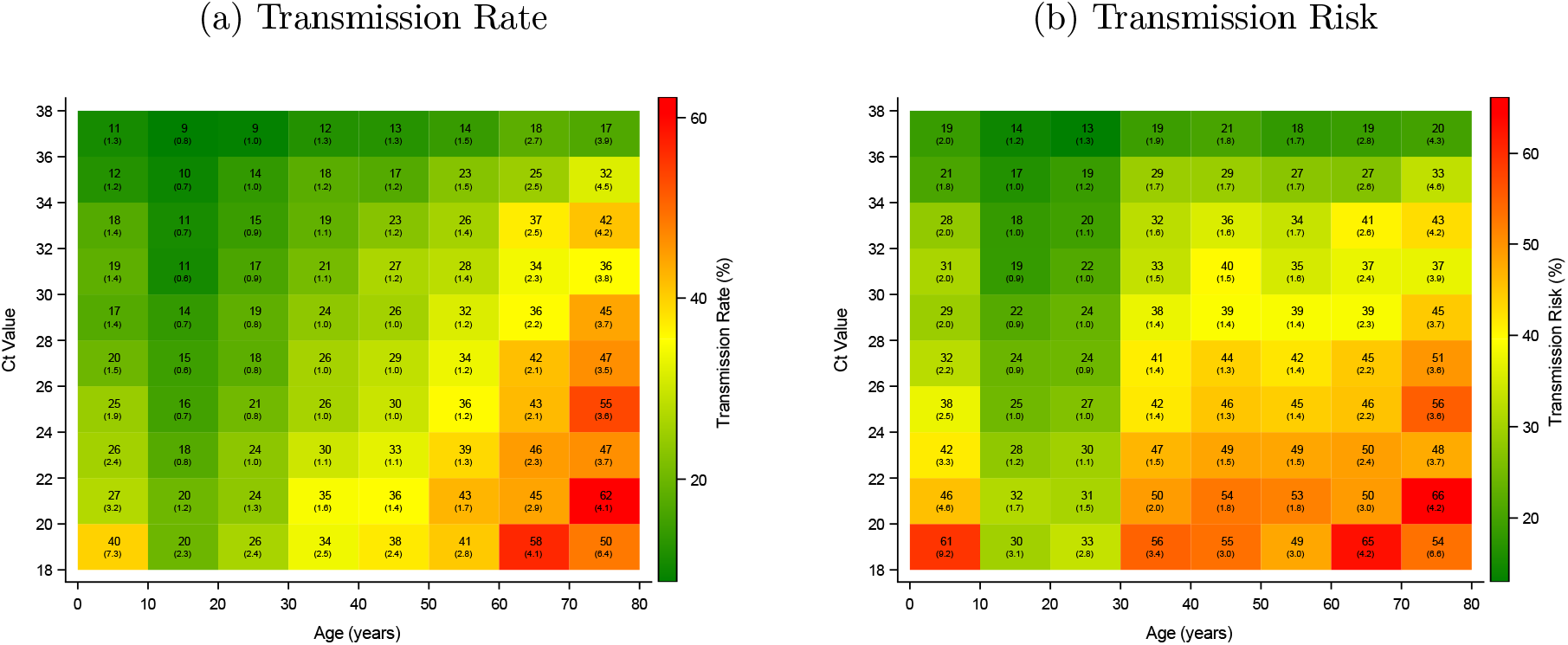
Association between age, Ct value, and transmissibility Notes: This figure shows the association between age, sample Ct value and transmissibility. The transmission rate describes the proportion of potential secondary cases within the household that were infected. The transmission risk describes the proportion of infected primary cases that infected at least one secondary case. For example, a primary case aged 30-40 years with a sample Ct value of 26-28 has a transmission rate of 26%, with a standard error of 1.0 (panel a). The transmission risk for this group was 41 with a standard error of 1.4 (panel b). Standard errors clustered on the household level in parenthesis.

Tables 2 and 3 provide odds ratio estimates for the transmissibility from a univariable and a multivariable regression analysis. Primary cases with a Ct value of 18 were approximately four times more transmissible compared with primary cases with a Ct value of 38. Children aged 0-5 years were approximately two times more transmissible compared with persons aged 15-20 years; whereas adults aged 70-75 years were 4-5 times more transmissible compared with persons ages 15-20 years. The transmission risk increased with the number of potential secondary cases, i.e., household size. The transmission rate was significantly higher for two person households compared to 3-6 person households.

**Table 2:** Odds ratio for transmission rate

|  | Univariable |  | Multivariable |  |
| --- | --- | --- | --- | --- |
|  | Odds Ratio | 95% CI | Odds Ratio | 95% CI |
| **Ct Value** |  |  |  |  |
| 18-20 | 1.00 | (.) | 1.00 | (.) |
| 20-22 | 0.89 | (0.83-0.96) | 0.95 | (0.95-0.95) |
| 22-24 | 0.76 | (0.71-0.81) | 0.83 | (0.83-0.83) |
| 24-26 | 0.65 | (0.61-0.69) | 0.72 | (0.72-0.72) |
| 26-28 | 0.59 | (0.55-0.63) | 0.66 | (0.66-0.66) |
| 28-30 | 0.53 | (0.50-0.57) | 0.59 | (0.59-0.59) |
| 30-32 | 0.46 | (0.43-0.50) | 0.52 | (0.52-0.52) |
| 32-34 | 0.43 | (0.40-0.47) | 0.49 | (0.49-0.49) |
| 34-36 | 0.35 | (0.32-0.38) | 0.39 | (0.39-0.39) |
| 36-38 | 0.24 | (0.22-0.26) | 0.26 | (0.25-0.26) |
| **Age** |  |  |  |  |
| 0 - 5 | 1.78 | (1.55-2.04) | 2.11 | (1.83-2.43) |
| 5 - 10 | 1.44 | (1.31-1.59) | 1.63 | (1.48-1.80) |
| 10 - 15 | 1.27 | (1.17-1.38) | 1.33 | (1.22-1.44) |
| 15 - 20 | 1.00 | (.) | 1.00 | (.) |
| 20 - 25 | 1.30 | (1.20-1.41) | 1.18 | (1.09-1.28) |
| 25 - 30 | 1.89 | (1.75-2.04) | 1.70 | (1.57-1.83) |
| 30 - 35 | 2.15 | (1.99-2.33) | 2.06 | (1.91-2.23) |
| 35 - 40 | 2.32 | (2.14-2.50) | 2.27 | (2.10-2.45) |
| 40 - 45 | 2.50 | (2.32-2.69) | 2.44 | (2.26-2.63) |
| 45 - 50 | 2.65 | (2.46-2.86) | 2.53 | (2.35-2.73) |
| 50 - 55 | 3.08 | (2.86-3.32) | 2.83 | (2.62-3.06) |
| 55 - 60 | 3.49 | (3.22-3.79) | 3.05 | (2.81-3.32) |
| 60 - 65 | 4.15 | (3.77-4.56) | 3.51 | (3.18-3.87) |
| 65 - 70 | 4.66 | (4.12-5.27) | 3.90 | (3.44-4.43) |
| 70 - 75 | 5.05 | (4.40-5.80) | 4.13 | (3.58-4.76) |
| 75 - 80 | 6.65 | (5.55-7.97) | 5.49 | (4.57-6.60) |
| 80 - 85 | 7.14 | (5.17-9.87) | 5.93 | (4.29-8.20) |
| 85 - 90 | 6.83 | (3.16-14.76) | 5.60 | (2.65-11.82) |
| **Sex** |  |  |  |  |
| Male | 1.00 | (.) | 1.00 | (.) |
| Female | 1.00 | (0.97-1.03) | 1.01 | (0.98-1.05) |
| **Household Size** |  |  |  |  |
| 2 | 1.00 | (.) | 1.00 | (.) |
| 3 | 0.63 | (0.61-0.66) | 0.77 | (0.73-0.80) |
| 4 | 0.63 | (0.60-0.66) | 0.79 | (0.75-0.83) |
| 5 | 0.61 | (0.58-0.64) | 0.79 | (0.75-0.84) |
| 6 | 0.58 | (0.54-0.63) | 0.75 | (0.69-0.81) |
| Observations |  | 139,882 |  | 139,882 |
| Households |  | 63,657 |  | 63,657 |

Notes: This table provides regression estimates for the odds ratio of a univariable and a multivariable model. The transmission rate describes the proportion of potential secondary cases within the household that were infected. 95% CI = 95% confidence intervals clustered on the household level.

**Table 3:** Odds ratio for transmission risk

|  | Univariable |  | Multivariable |  |
| --- | --- | --- | --- | --- |
|  | Odds Ratio | 95% CI | Odds Ratio | 95% CI |
| **Ct Value** |  |  |  |  |
| 18-20 | 1.00 | (.) | 1.00 | (.) |
| 20-22 | 0.89 | (0.79-1.01) | 0.93 | (0.84-1.03) |
| 22-24 | 0.76 | (0.68-0.84) | 0.81 | (0.74-0.89) |
| 24-26 | 0.64 | (0.58-0.71) | 0.69 | (0.64-0.76) |
| 26-28 | 0.59 | (0.53-0.65) | 0.64 | (0.59-0.70) |
| 28-30 | 0.51 | (0.46-0.57) | 0.56 | (0.51-0.62) |
| 30-32 | 0.45 | (0.41-0.50) | 0.50 | (0.45-0.55) |
| 32-34 | 0.43 | (0.39-0.48) | 0.46 | (0.42-0.51) |
| 34-36 | 0.34 | (0.30-0.38) | 0.36 | (0.32-0.40) |
| 36-38 | 0.23 | (0.20-0.26) | 0.24 | (0.21-0.27) |
| **Age** |  |  |  |  |
| 0 - 5 | 1.95 | (1.69-2.25) | 2.17 | (1.87-2.51) |
| 5 - 10 | 1.61 | (1.46-1.77) | 1.66 | (1.50-1.83) |
| 10 - 15 | 1.43 | (1.32-1.55) | 1.37 | (1.26-1.49) |
| 15 - 20 | 1.00 | (.) | 1.00 | (.) |
| 20 - 25 | 1.02 | (0.95-1.11) | 1.25 | (1.15-1.35) |
| 25 - 30 | 1.63 | (1.51-1.76) | 2.00 | (1.85-2.16) |
| 30 - 35 | 2.34 | (2.17-2.54) | 2.51 | (2.32-2.72) |
| 35 - 40 | 2.81 | (2.59-3.04) | 2.74 | (2.53-2.98) |
| 40 - 45 | 2.92 | (2.71-3.16) | 2.84 | (2.63-3.08) |
| 45 - 50 | 2.83 | (2.62-3.06) | 2.95 | (2.73-3.19) |
| 50 - 55 | 2.74 | (2.54-2.96) | 3.28 | (3.03-3.56) |
| 55 - 60 | 2.61 | (2.41-2.84) | 3.51 | (3.22-3.82) |
| 60 - 65 | 2.83 | (2.57-3.12) | 4.02 | (3.63-4.45) |
| 65 - 70 | 3.04 | (2.68-3.44) | 4.49 | (3.94-5.11) |
| 70 - 75 | 3.25 | (2.82-3.73) | 4.78 | (4.13-5.54) |
| 75 - 80 | 4.20 | (3.50-5.04) | 6.39 | (5.29-7.72) |
| 80 - 85 | 4.42 | (3.18-6.13) | 6.61 | (4.74-9.22) |
| 85 - 90 | 4.02 | (1.86-8.68) | 5.74 | (2.59-12.74) |
| **Sex** |  |  |  |  |
| Male | 1.00 | (.) | 1.00 | (.) |
| Female | 0.99 | (0.96-1.03) | 1.03 | (1.00-1.07) |
| **Household Size** |  |  |  |  |
| 2 | 1.00 | (.) | 1.00 | (.) |
| 3 | 1.05 | (1.00-1.10) | 1.32 | (1.26-1.39) |
| 4 | 1.33 | (1.27-1.38) | 1.77 | (1.68-1.86) |
| 5 | 1.52 | (1.44-1.61) | 2.16 | (2.03-2.30) |
| 6 | 1.71 | (1.57-1.87) | 2.41 | (2.19-2.64) |
| Observations |  | 139,882 |  | 139,882 |
| Households |  | 63,657 |  | 63,657 |

Notes: The transmission risk describes the proportion of infected primary cases that infected at least one secondary case. 95% CI = 95% confidence intervals clustered on the household level.

### 3.3 Sensitivity analyses

The trends of the age structured transmissibility were similar across males and females (Figure S3). Boys (<15 years) had a significantly higher transmission risk than girls. We found the same overall transmission pattern when stratifying by Ct value quartiles instead of the median, underlining the robustness of dichotomization by the median (Figure S4). We found that primary cases tested at hospital test facilities generally had slightly higher transmissibility (Figure S5).

Furthermore, when estimating the transmission rate by the interaction of the age of the primary case and the age of the potential secondary cases, we found the same overall transmission pattern across Ct value quartiles (Figure S6). Generally, primary cases with lower Ct values had a higher transmission rate. Also, adult primary cases (>30 years) generally had a higher transmission rate to persons around their own age.

Lastly, in Appendix C, the results of the sensitivity analyses for varying the inclusion criteria of secondary cases are provided. When we excluded more cases identified temporally close to the primary case, we found that the association between Ct values and transmissibility decreased, although the patterns remained the same and were still significant (Figure S7). That is, when we excluded secondary cases identified in the days right after the primary case was identified, we generally exclude secondary cases associated with primary cases that had low Ct values. This is further illustrated in the sensitivity analysis of Figure 2, where the proportion of secondary cases associated with Ct values ≥30 increased from 34% to 38%, when we only included secondary cases identified on days 4-14 (Figure S8). The differences in the age structured transmissibility across primary cases with a Ct value below and above the median were reduced when we excluded secondary cases identified shortly after the primary case was identified (Figure S11 and S12).

## 4 Discussion and Conclusion

In this nationwide study, we found an approximately linear association between Ct value and transmissibility. Under the assumption that the sample Ct value reflects the viral load when transmission occurred, this suggests that cases with a higher viral load are more infectious than cases with a lower viral load. This was expected, because a high viral load implies that there are more virus particles in the sample, and hence the possibility of a larger inoculum. This finding corroborates previous studies (Marks et al., 2021; Lee et al., 2021a,b), however, the present study also highlights that there is no obvious cut-off for Ct values to eliminate transmissibility. Importantly, we found a considerable transmissibility for cases with high Ct values, e.g., primary cases with a Ct value of 38 had a transmission risk of 13% and a transmission rate of 9% within the household. This is similar to a previous study where virus could be isolated from 8.3% of samples with Ct >35 (Singanayagam et al., 2020) and does not corroborate previous studies that have argued that cases with a Ct value above a certain cut-off are not contagious. A Ct value cut-off of 30, or even lower, has been suggested (La Scola et al., 2020; Bullard et al., 2020; Brown et al., 2020; Prince-Guerra et al., 2021), as this Ct value corresponds to the limit of detection of virus cultures in Vero cells and antigen tests. Whereas this study does not test whether cases a low viral load are contagious (as discussed below, viral load changes over times and obtained sample Ct values depend on technical issues), it shows that choosing a cut-off of a Ct≤30 for infectiousness instead of Ct≤38 would have missed transmission to 34% of the secondary cases in this study of household transmission.

Another main result from this study is that age was strongly associated with transmissibility. We found that younger children were more transmissible compared with older children—both for transmission rate and transmission risk. For adults (≥20 years), we found an over overall positive association between age and transmissibility. However, the transmission risk plateaued for persons aged 30-60 years old. The two associations with transmissibility—age and viral load—were also found when investigating them together in the same analysis (Figure S6 and Table 2 and 3).

This age-related transmissibility pattern could be driven by the susceptibility of the potential secondary cases, as people tend to live with their partner, who is around their own age, and parents live with their children (Lyngse et al., 2020; Madewell et al., 2020). We investigated this and did find that primary cases tend to infect other people around the same age within the household. Moreover, it is likely that the higher transmission from smaller children compared with teenagers is related to the fact that children below 10 years, to a larger degree, need parental contact and nursing compared with teenagers who are more independent and can self-isolate more efficiently. The age-related transmissibility among adults may also be due to increased viral exhalation by age (Edwards et al., 2021) or that the immunological response decreases with age (Long et al., 2020). Further research is needed to clarify this. Heald-Sargent et al. (2020) studied the viral load in 46 cases aged 0-5 years, 51 cases aged 5-17 years, and 48 cases aged 18-65 years. They found indications of high viral loads in children and speculated that they could be a main driver of the epidemic. Our results, which was based on a large sample, contradict this, as we did not find any association between the distribution of Ct values and age (Table S4). To calculate the distribution of Ct values across age groups, it is necessary to include a sample comprising all age groups, as in the present study.

During an infection, the viral load is low shortly after exposure, increases over the infection, peaks around the onset of symptoms, and decreases later on (He et al., 2020). Hence, the Ct value is sensitive to the timing of the test, e.g., when a case is pre-symptomatic compared to later when a case is symptomatic. Furthermore, there is a large variation in both severity of symptoms and infectivity across persons. One study found an indication of lower viral loads in asymptomatic persons (Zhou et al., 2020), while another found no differences across symptomatic and asymptomatic cases (Long et al., 2020). Additionally, the test results depend on the quality of the sampling as well as the assay, i.e., the chosen primers, probe and other reagents, which determines the accuracy of the test, making comparisons across laboratories difficult. In this study, the same methodology, set of primers, and probe were used throughout the whole study period, making the Ct values comparable. Theoretically, it could be possible that the finding of a considerable transmissibility for high Ct values (low viral load) were due to sampling late in the infection and secondary cases were actually infected earlier, when the primary case had a lower Ct value (high viral load). If this was the case, we would find a shift in the estimates for higher Ct values, when we exclude secondary cases identified in the days shortly after identification of the primary case. However, even in the extreme case where we only include secondary cases that tested positive 4-14 days after the primary case, we still find a 6% transmission rate and 8% transmission risk for cases with a Ct value of 38, and 38% of all secondary cases come from cases with a Ct≥ 30. Thus, our results show that primary cases with low viral loads judged by the sample Ct value had a transmissibility significantly above zero, even when we only included secondary cases identified on days 4-14. This indicates that primary cases identified at a time with a low viral load were able to generate secondary cases (Figure S7). Despite the variations mentioned above, we found a clear association between Ct values and transmission risk, emphasizing the importance of this association.

There are several strengths in the present study. Our estimates benefit from a large sample size and a systematic selection of potential secondary cases. We included all household members as potential secondary cases (unconditional on them being contacted by the official contact tracing system), of which 82% were tested within 1-14 days of the primary case. It can be argued that some of the secondary cases might represent cases infected outside of the household. Lyngse et al. (2021) used genomic data (whole genome sequencing) to validate the same methods as used in the present study. They found an intra-household correlation of lineages between primary and positive secondary cases of 96-99%, indicating that most secondary cases were infected with the same lineage as the primary case.

Some limitations apply to this study. We defined the primary cases as the first positive test within a household and all other people living in the same household as potential secondary cases. We defined all secondary cases as those testing positive 1-14 days after the primary case. However, some of these co-primary and secondary cases may be misclassified, e.g., if they were infected earlier but not diagnosed, because they were pre- or asymptomatic. Including secondary cases found >14 days after the primary case could result in misclassification of secondary cases being either tertiary cases or having somewhere else as the source of secondary infections. For a thorough discussion of the definition of co-primary cases, see Lyngse et al. (2020).

Our results are important for public health. Optimal contact tracing naturally has to prioritize the order of new cases and their contacts. Our results suggest that contact tracing should prioritize cases according to Ct values and age. However, it should be noted that primary cases with high Ct values (low viral loads) had a transmissibility significantly above zero and that there is no obvious cut-off of Ct values to eliminate transmissibility. Furthermore, households can potentially serve as a transmission bridge by creating connections between otherwise separate domains, such as schools and workplaces. Lastly, our results underline the importance of quick identification and isolation of cases, especially since non-pharmaceutical interventions are difficult to apply in the household domain.

In conclusion, although primary cases with high viral loads (low Ct values) were associated with higher SARS-CoV-2 transmissibility, we found no obvious cut-off for Ct values to eliminate transmissibility. Even in households where the samples from primary cases had a high Ct value (low viral load), transmission occurred, and these households accounted for a substantial number of secondary cases. We further found a clear increasing association between age and transmissibility. These results are important for public health, as they suggest that contact tracing should prioritize cases according to Ct values and age, and underline the importance of quick identification and isolation of cases. Furthermore, households can serve as a transmission bridge by creating connections between otherwise separate domains.

## Data Availability

Data access can be granted through application to the Danish Health Data Authority.

## Appendix A

### Summary statistics

**Table S1:** Summary statistics: RT-PCR SARS-CoV-2 tests

|  | TCDK |  |  | Hospitals |  |  | Total |  |  |
| --- | --- | --- | --- | --- | --- | --- | --- | --- | --- |
|  | N | % | % | N | % | % | N | % | % |
|  | Obs. | Tot. | Ind. | Obs. | Tot. | Ind. | Obs. | Tot. | Ind. |
| **All tests** |  |  |  |  |  |  |  |  |  |
| Tests | 9,347,218 | 75 |  | 3,111,731 | 25 |  | 12,458,949 | 100 |  |
| Positive tests | 145,891 | 70 |  | 63,725 | 30 |  | 209,616 | 100 |  |
| Individuals | 3,327,337 | 84 |  | 1,601,518 | 40 |  | 3,968,337 | 100 |  |
| **Primary Cases** |  |  |  |  |  |  |  |  |  |
| Individuals | 63,937 | 73 |  | 23,973 | 27 |  | 87,910 | 100 |  |
| With Ct value | 63,657 |  | 100 | 0 |  | 0 | 63,657 |  | 72 |
| **Potential Secondary Cases** |  |  |  |  |  |  |  |  |  |
| Individuals | 140,470 | 74 |  | 50,489 | 26 |  | 190,959 | 100 |  |
| Tested | 115,633 |  | 82 | 41,755 |  | 83 | 157,388 |  | 82 |
| Tested positive | 29,863 |  | 21 | 12,297 |  | 24 | 42,160 |  | 22 |

Notes: This table provides summary statistics for all SARS-CoV-2 RT-PCR tests taken from August 25, 2020 to February 10, 2021, stratified by place of testing. Approximately, 75% of all tests were taken in TCDK and 25% in hospitals. Similarly, 73% of all primary cases were identified in TCDK and 23% in hospitals. For primary cases identified in TCDK, 99.6% had a Ct value registered on the sample, whereas we had no Ct values for primary cases identified in hospitals. N Obs. = number of observations. % Tot. = percentage of total. % Ind. = percentage of individuals.

**Table S2:** Summary statistics: Sex, age, household size

|  | **Primary**<br>**Cases** | **Potential**<br>**Secondary**<br>**Cases** | **Positive**<br>**Secondary**<br>**Cases** | **Attack**<br>**Rate**<br>**(%)** |
| --- | --- | --- | --- | --- |
| **Total** | 63,657 | 139,882 | 29,739 | 21 |
| **Sex** |  |  |  |  |
| Male | 32,256 | 70,849 | 14,646 | 21 |
| Female | 31,401 | 69,002 | 15,090 | 22 |
| **Age** |  |  |  |  |
| 0 - 5 | 977 | 13,025 | 2,041 | 16 |
| 5 - 10 | 2,734 | 11,990 | 2,692 | 22 |
| 10 - 15 | 5,113 | 14,837 | 3,156 | 21 |
| 15 - 20 | 9,167 | 14,440 | 2,691 | 19 |
| 20 - 25 | 7,498 | 11,426 | 1,877 | 16 |
| 25 - 30 | 5,934 | 8,711 | 1,828 | 21 |
| 30 - 35 | 4,704 | 7,214 | 1,760 | 24 |
| 35 - 40 | 4,110 | 7,820 | 1,841 | 24 |
| 40 - 45 | 4,568 | 11,383 | 2,421 | 21 |
| 45 - 50 | 4,825 | 13,471 | 2,699 | 20 |
| 50 - 55 | 4,777 | 11,106 | 2,440 | 22 |
| 55 - 60 | 4,004 | 6,689 | 1,710 | 26 |
| 60 - 65 | 2,422 | 3,524 | 1,096 | 31 |
| 65 - 70 | 1,228 | 1,851 | 623 | 34 |
| 70 - 75 | 924 | 1,346 | 498 | 37 |
| 75 - 80 | 499 | 675 | 239 | 35 |
| 80 - 85 | 147 | 248 | 102 | 41 |
| 85 - 90 | 26 | 72 | 19 | 26 |
| 90 - 95 | - | 18 | <5 | - |
| >95 | - | 5 | <5 | - |
| **Household Size** |  |  |  |  |
| 2 | 21,830 | 21,830 | 6,345 | 29 |
| 3 | 15,226 | 29,375 | 5,948 | 20 |
| 4 | 16,773 | 48,532 | 9,725 | 20 |
| 5 | 7,528 | 29,066 | 5,642 | 19 |
| 6 | 2,300 | 11,079 | 2,079 | 19 |

Notes: This table provides summary statistics for the number of primary cases, potential secondary cases, positive secondary cases, and attack rates in the study, stratified by sex, age and household sizes. For example, in the 0-5 year age group, there are 977 primary cases, 13,025 potential secondary cases, and 2,041 positive secondary cases. This implies an attack rate for 0-5 year olds of 16% (2,041/13,025). For 31 potential secondary cases, age and sex were not observed.

**Table S3:** Summary statistics: Ct values of primary cases

| **Ct value** | **Primary Cases** | **Potential Secondary Cases** | **Positive Secondary Cases** | **Attack Rate (%)** |
| --- | --- | --- | --- | --- |
| <18 | 155 | 340 | 72 | 21 |
| 18 | 205 | 415 | 177 | 43 |
| 19 | 488 | 1,001 | 357 | 36 |
| 20 | 998 | 2,141 | 674 | 31 |
| 21 | 1,710 | 3,668 | 1,149 | 31 |
| 22 | 2,618 | 5,710 | 1,689 | 30 |
| 23 | 3,371 | 7,251 | 2,056 | 28 |
| 24 | 3,918 | 8,511 | 2,216 | 26 |
| 25 | 4,407 | 9,674 | 2,360 | 24 |
| 26 | 4,761 | 10,353 | 2,443 | 24 |
| 27 | 4,865 | 10,474 | 2,361 | 23 |
| 28 | 4,668 | 10,287 | 2,213 | 22 |
| 29 | 4,481 | 9,670 | 1,965 | 20 |
| 30 | 4,169 | 9,204 | 1,883 | 20 |
| 31 | 3,875 | 8,659 | 1,599 | 18 |
| 32 | 3,516 | 7,734 | 1,428 | 18 |
| 33 | 3,273 | 7,384 | 1,282 | 17 |
| 34 | 3,108 | 7,051 | 1,198 | 17 |
| 35 | 2,801 | 6,338 | 945 | 15 |
| 36 | 2,596 | 5,815 | 798 | 14 |
| 37 | 2,511 | 5,689 | 669 | 12 |
| 38 | 1,163 | 2,513 | 205 | 8 |
| Total | 63,657 | 139,882 | 29,739 | 21 |

Notes: This table provides summary statistics for primary cases according to the Ct value of their first positive test, and their associated potential secondary cases and positive secondary cases, as well as the attack rate. For example, there are 1,163 primary cases with a Ct value of 38. They live with 2,513 potential secondary cases, of which 205 are positive secondary cases. This implies an attack rate of 8% (205/2,513). An RT-PCR test is positive if the Ct value is $\leq 38$ .

**Figure S1:**
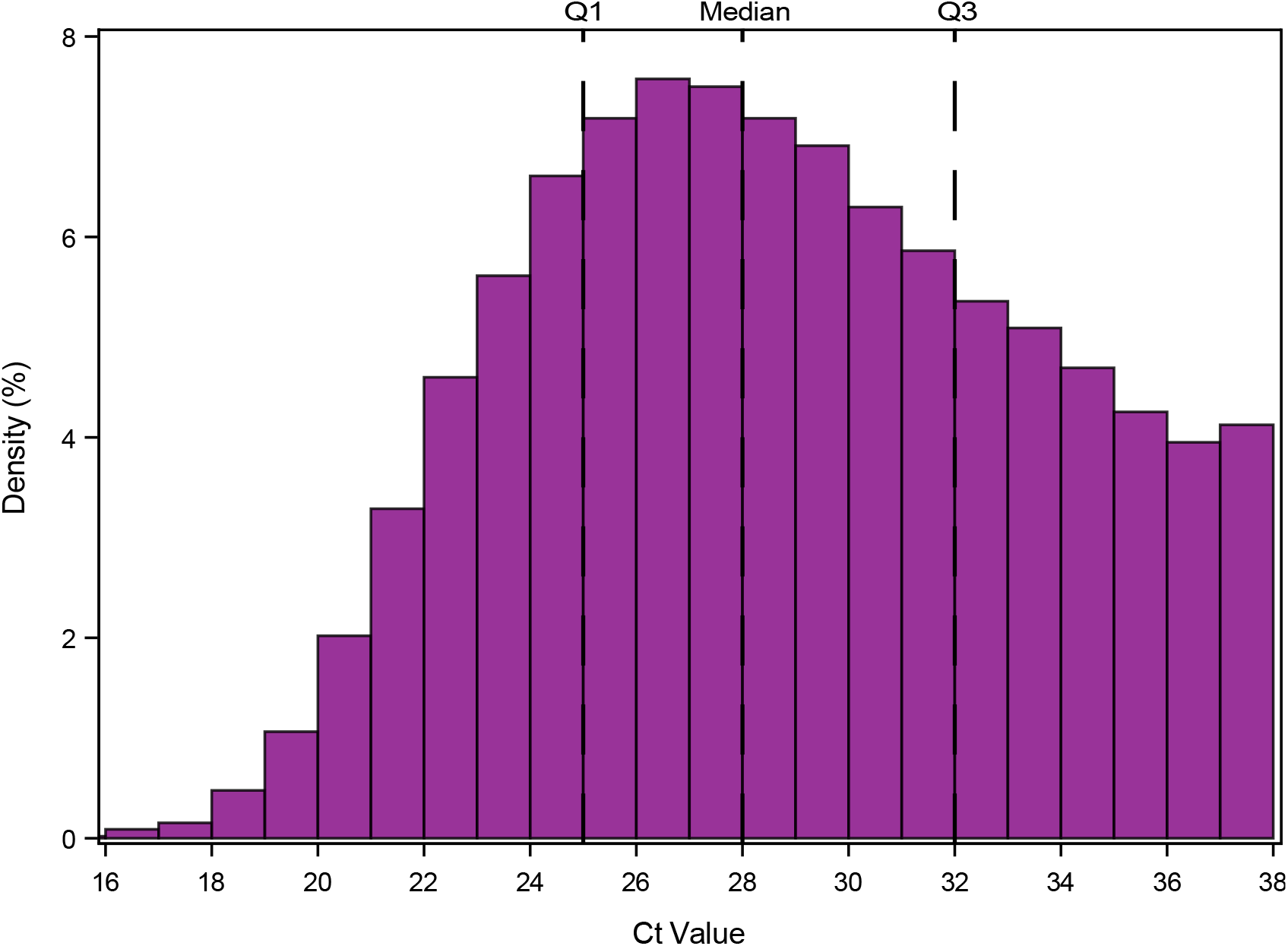
Distribution of Ct values for primary cases Notes: This figure shows the distribution of Ct values for primary cases in the study. Only Ct values of the first positive test of each primary case are shown. Q1 = 1st quartile (25th percentile, P25), Q3 = 3rd quartile (75th percentile, P75). An RT-PCR test is positive if the Ct value is ≤38.

**Figure S2:**
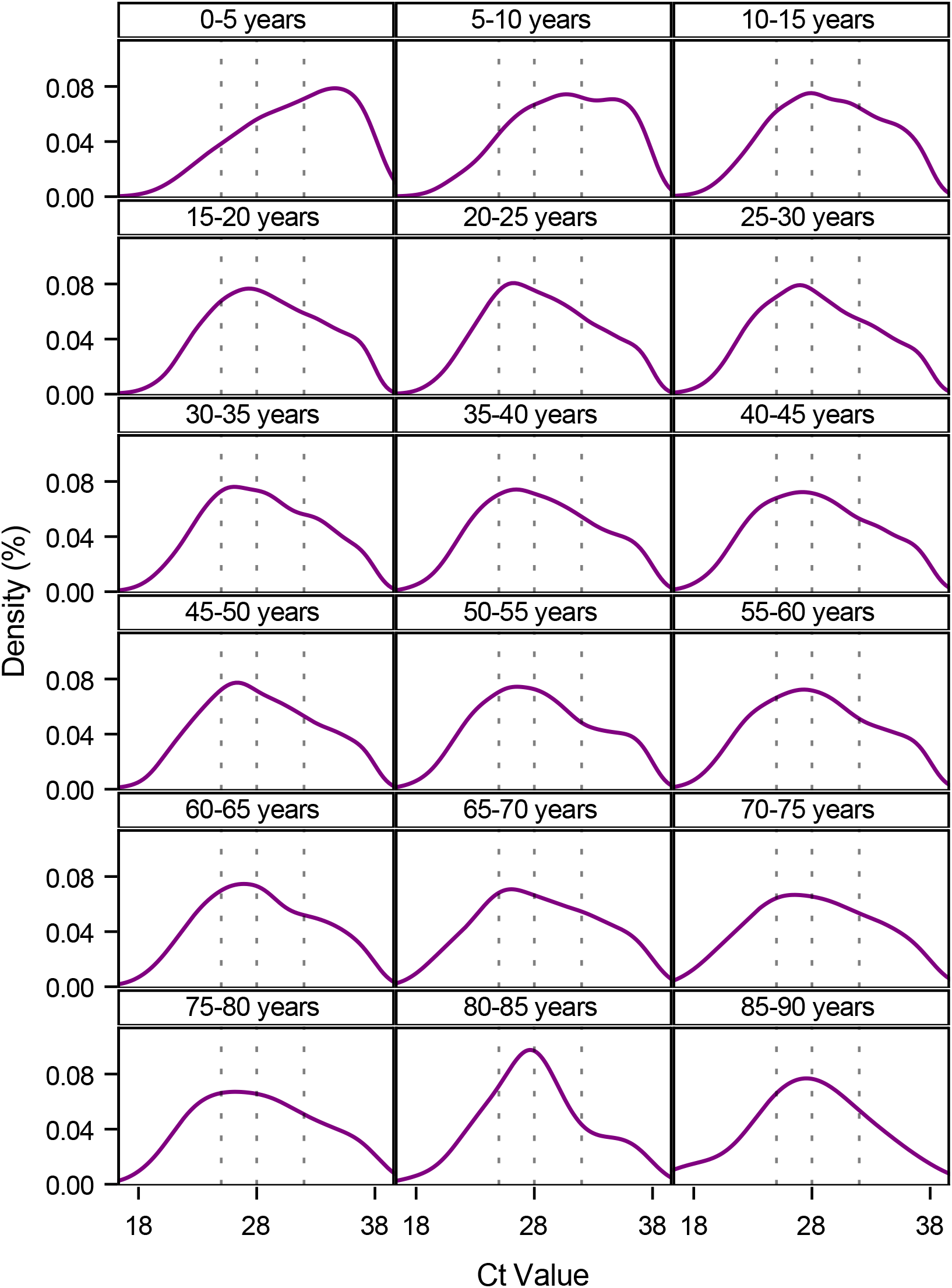
Distribution of Ct values for primary cases stratified by age Notes: This figure shows the kernel density plots of the Ct values for primary cases stratified by five year age groups. Only Ct values of the first positive test of primary cases are shown. The vertical dotted reference lines are the 1st quartile, the median, and 3rd quartile of the total population. The number of primary cases for each group as well as distributional figures can be found in Table S4. An RT-PCR test is positive if the Ct value is ≤38.

**Table S4:** Distribution of Ct values for primary cases

|  | **P5** | **P10** | **P25** | **P50** | **P75** | **P90** | **P95** | **Mean** | **Observations** |
| --- | --- | --- | --- | --- | --- | --- | --- | --- | --- |
| **Total** | 21 | 23 | 25 | 28 | 32 | 35 | 37 | 29 | 63,657 |
| **Sex** |  |  |  |  |  |  |  |  |  |
| Male | 21 | 22 | 25 | 28 | 32 | 35 | 37 | 28 | 32,256 |
| Female | 22 | 23 | 25 | 29 | 33 | 36 | 37 | 29 | 31,401 |
| **Age** |  |  |  |  |  |  |  |  |  |
| 0 - 5 | 23 | 24 | 28 | 32 | 35 | 37 | 38 | 31 | 977 |
| 5 - 10 | 23 | 25 | 27 | 31 | 34 | 36 | 37 | 31 | 2,734 |
| 10 - 15 | 22 | 23 | 26 | 29 | 33 | 36 | 37 | 29 | 5,113 |
| 15 - 20 | 22 | 23 | 25 | 29 | 33 | 35 | 37 | 29 | 9,167 |
| 20 - 25 | 22 | 23 | 25 | 28 | 32 | 35 | 37 | 29 | 7,498 |
| 25 - 30 | 21 | 22 | 25 | 28 | 32 | 35 | 37 | 28 | 5,934 |
| 30 - 35 | 21 | 23 | 25 | 28 | 32 | 35 | 36 | 29 | 4,704 |
| 35 - 40 | 21 | 22 | 25 | 28 | 32 | 35 | 37 | 28 | 4,110 |
| 40 - 45 | 21 | 22 | 25 | 28 | 32 | 35 | 37 | 28 | 4,568 |
| 45 - 50 | 21 | 22 | 25 | 28 | 32 | 35 | 37 | 28 | 4,825 |
| 50 - 55 | 21 | 22 | 25 | 28 | 32 | 36 | 37 | 28 | 4,777 |
| 55 - 60 | 21 | 22 | 25 | 28 | 32 | 35 | 37 | 28 | 4,004 |
| 60 - 65 | 21 | 22 | 25 | 28 | 32 | 35 | 37 | 28 | 2,422 |
| 65 - 70 | 20 | 22 | 25 | 28 | 32 | 35 | 37 | 28 | 1,228 |
| 70 - 75 | 20 | 22 | 24 | 28 | 32 | 35 | 37 | 28 | 924 |
| 75 - 80 | 21 | 22 | 24 | 28 | 32 | 36 | 37 | 28 | 499 |
| 80 - 85 | 21 | 23 | 25 | 28 | 31 | 35 | 36 | 28 | 147 |
| 85 - 90 | 19 | 22 | 25 | 28 | 31 | 34 | 36 | 28 | 26 |

Notes: This table provides summary statistics on the distribution of sample Ct values for primary cases. Only Ct values of the first positive test of primary cases are shown. An RT-PCR test is positive if the Ct value is $\leq 38$ . P5 = 5th percentile, P25 = 25th percentile (1st quartile), etc.

## Appendix B

### Sensitivity Analyses

**Figure S3:**
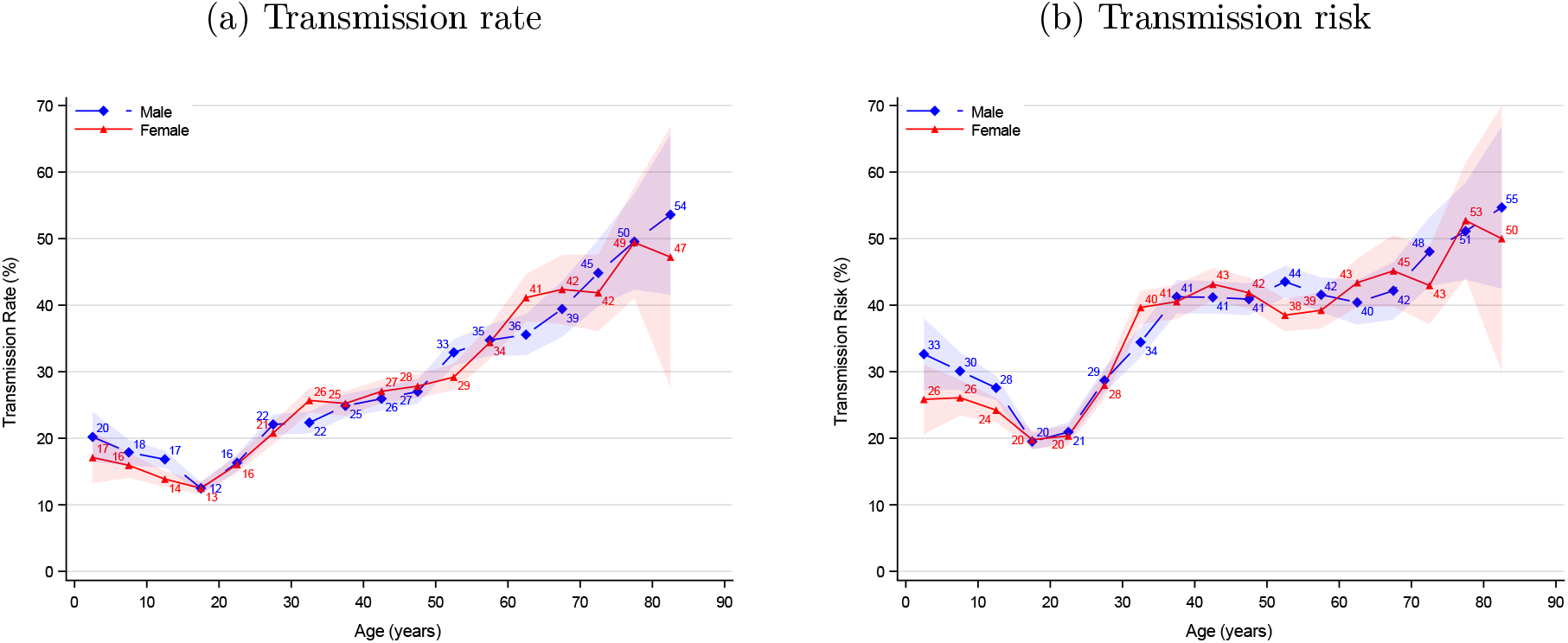
Association between age structured transmissibility and sex Notes: This figure shows the age structured transmissibility estimates stratified by sex: males (blue) and females (red). The shaded areas show the 95% confidence bands clustered on the household level.

**Figure S4:**
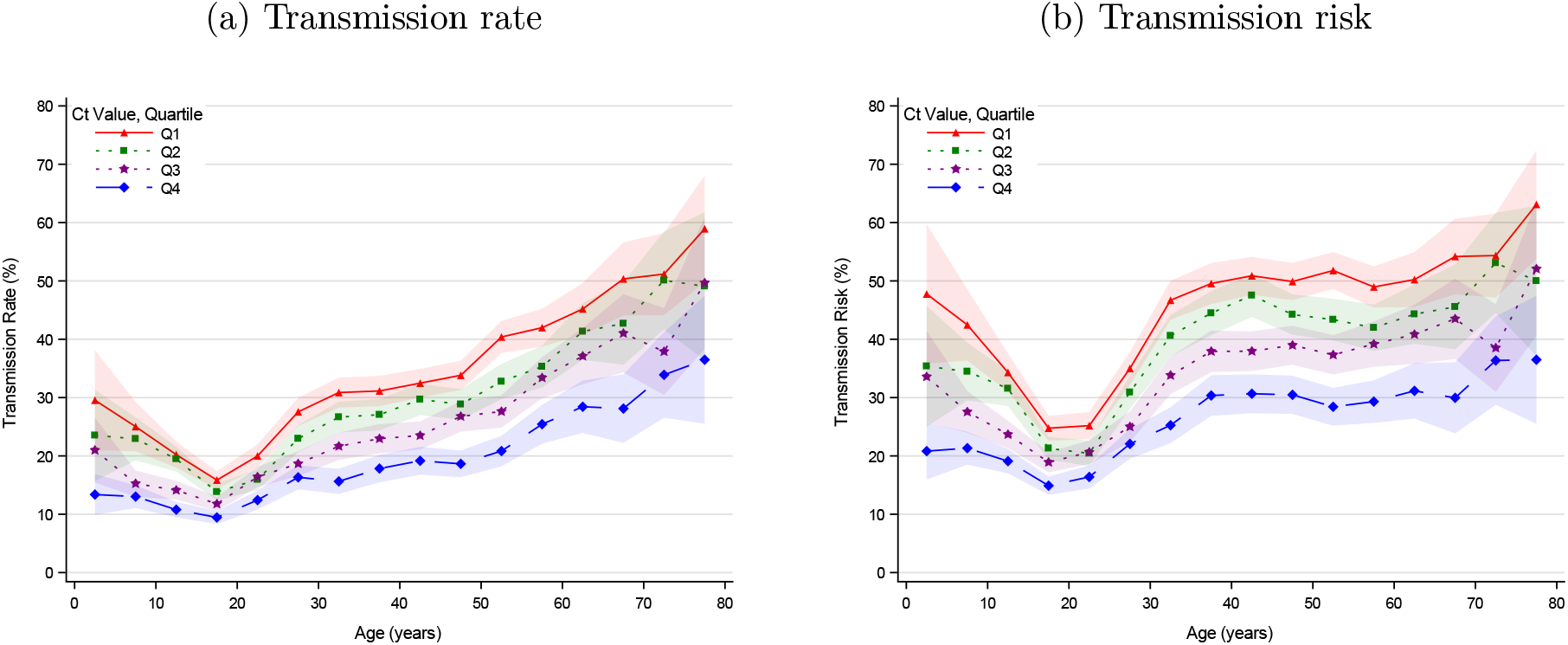
Association between Ct value (quartiles) and age structured transmissibility Notes: This figure shows the age structured transmissibility estimates stratified by sample Ct value quartiles. (Q1≤25, 25<Q2≤28, 28<Q3≤32, 32<Q4≤38.) The shaded areas show the 95% confidence bands clustered on the household level.

**Figure S5:**
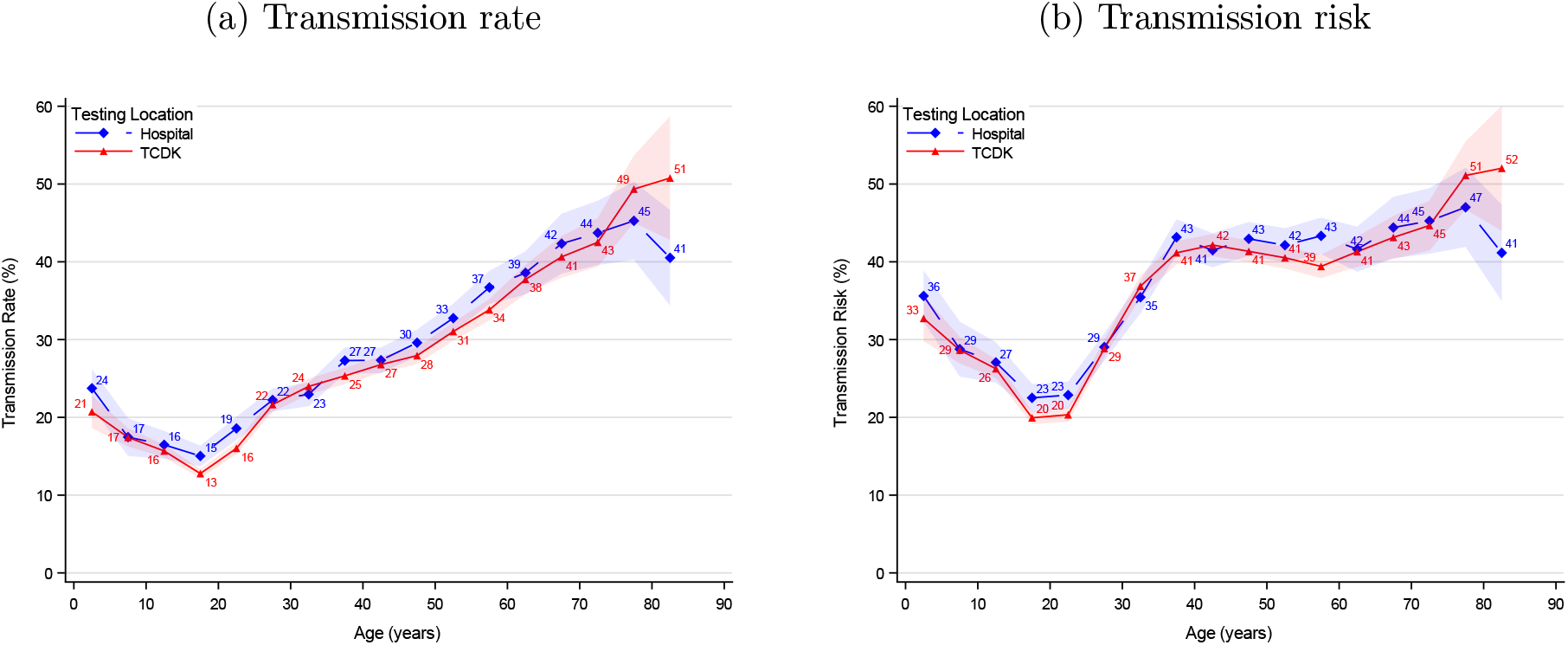
Association between age structured transmissibility by place of testing Notes: This figure shows the age structured transmissibility estimates stratified by place of testing: TCDK (red) and hospitals (blue). The shaded areas show the 95% confidence bands clustered on the household level.

**Figure S6:**
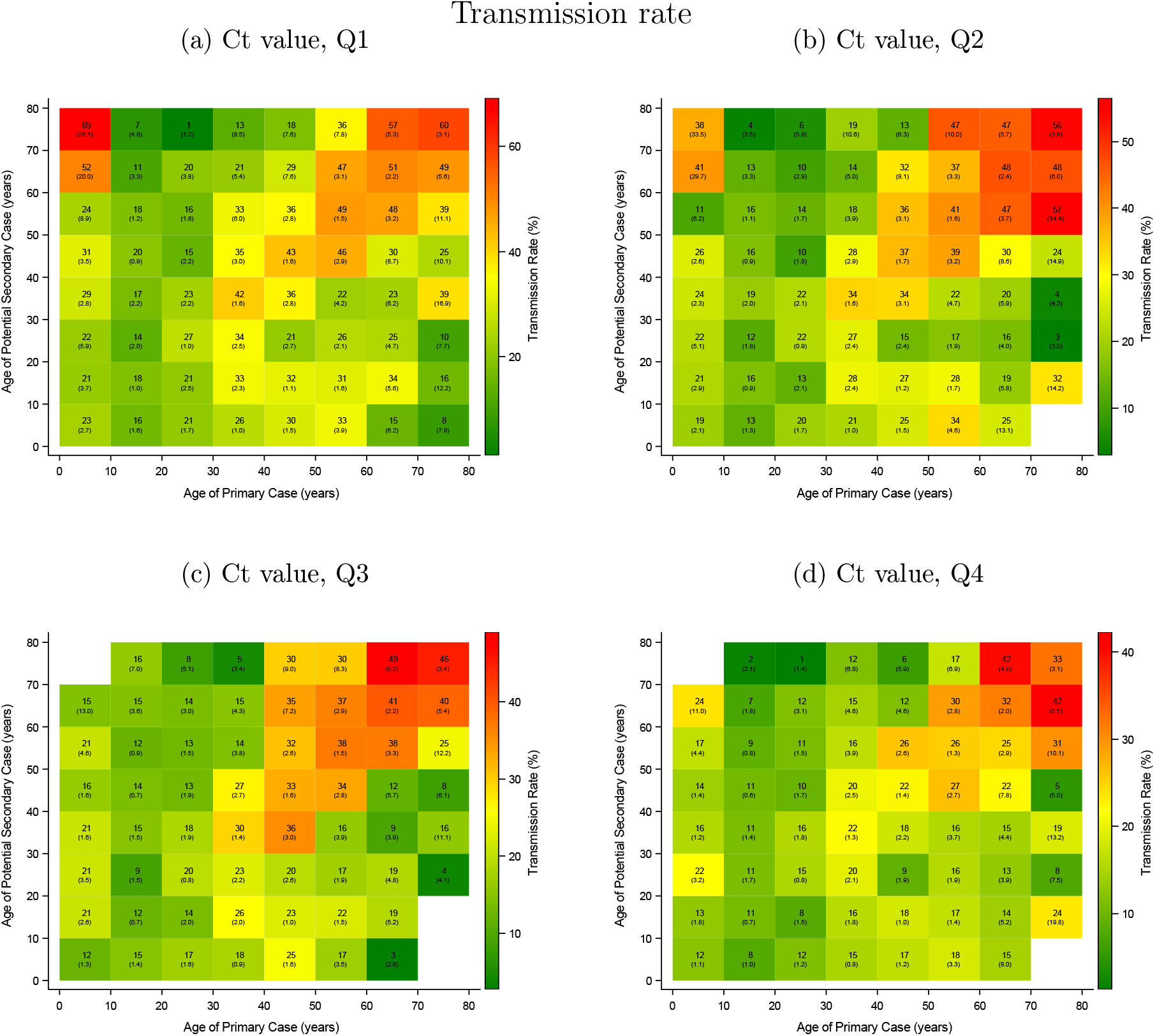
Association between age of primary case, age of potential secondary case, and transmission rate—stratified by Ct value quartile Notes: This figure shows the transmission rate from the interaction between age of the primary case and age of the potential secondary cases. Panel (a) shows the transmission rate for primary cases with sample Ct values in the lowest quartile, Q1. (Q1≤25, 25<Q2≤28, 28<Q3≤32, 32<Q4≤38.) Standard errors clustered on the household level in parentheses.

## Appendix C

### Sensitivity analyses for definition of secondary cases

In this appendix, we varied the inclusion criteria of secondary cases. In the main results, we included secondary cases that tested positive 1-14 days following the primary case. In this appendix, we vary the inclusion criteria for secondary cases. Panel a includes secondary cases that tested positive 1-14 days after the primary case, and is the same as the main results. Panel b includes secondary cases that tested positive 2-14 days after the primary case. Panel c includes secondary cases that tested positive 3-14 days after the primary case. Panel d includes secondary cases that tested positive 4-14 days after the primary case. Table S5 provides summary statistics on the number of primary cases, potential secondary cases and positive secondary cases for each of the four inclusion criteria.

**Table S5:** Summary statistics for sensitivity of secondary cases

|  | **I** | **II** | **III** | **IV** |
| --- | --- | --- | --- | --- |
| Days for including secondary cases | **1-14** | **2-14** | **3-14** | **4-14** |
| Primary cases | 63,657 | 62,422 | 60,224 | 58,805 |
| (%) | (100) | (98) | (95) | (92) |
| Potential secondary cases | 139,882 | 135,449 | 128,497 | 124,293 |
| (%) | (100) | (97) | (92) | (89) |
| Positive secondary cases | 29,739 | 25,306 | 18,354 | 14,150 |
| (%) | (100) | (85) | (62) | (48) |

Notes: This table provides summary statistics on the number of primary cases, potential secondary cases, and positive secondary cases depending on the inclusion criteria of secondary cases. Column I includes secondary cases that tested positive 1-14 days after the primary case and is the same as the main analyses. Column II includes secondary cases that tested positive 2-14 days after the primary case. Column III includes secondary cases that tested positive 3-14 days after the primary case. Column IV includes secondary cases that tested positive 4-14 days after the primary case.

**Figure S7:**
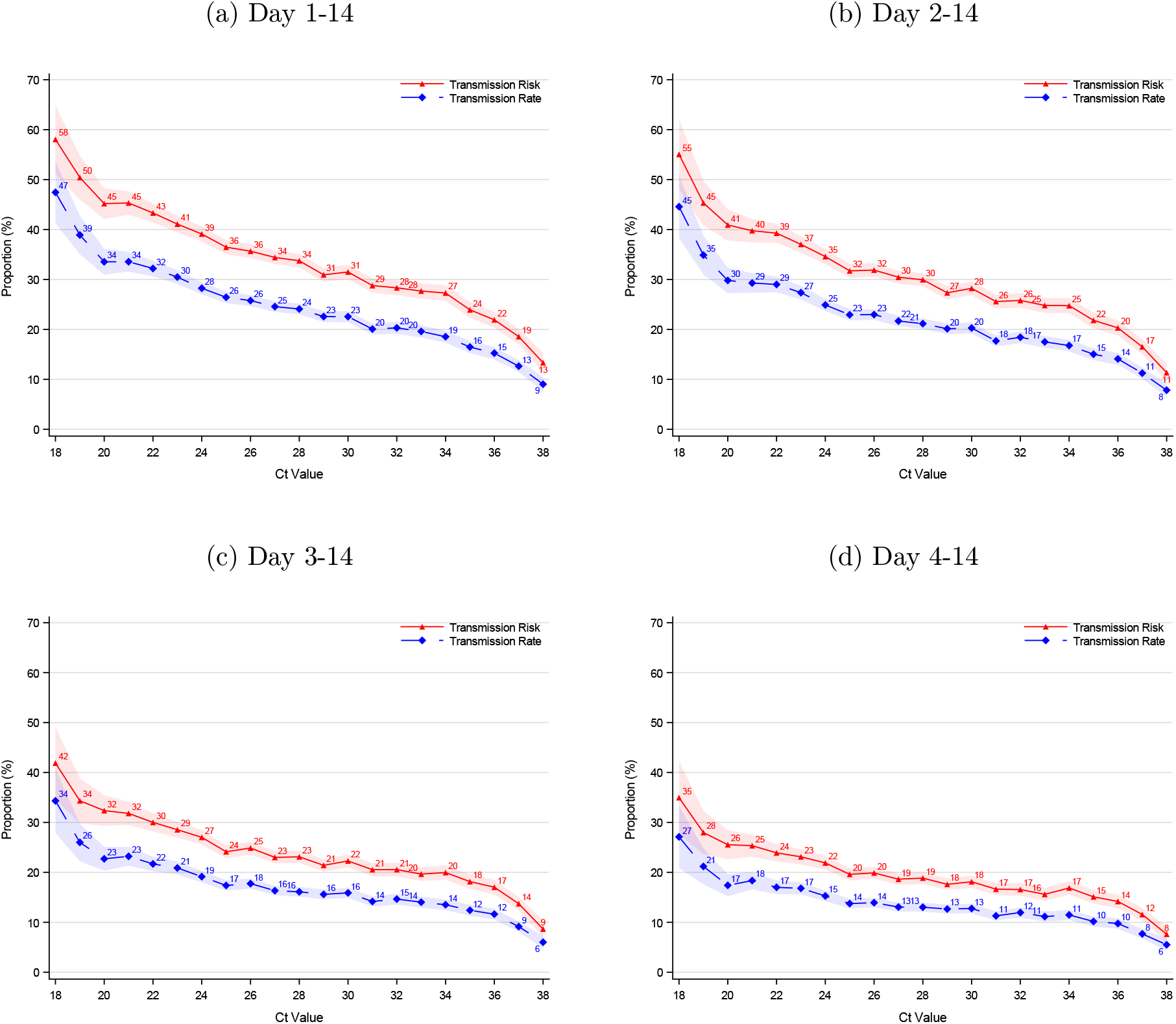
Sensitivity to definition of secondary cases: Association between Ct values and transmissibility Notes: This figure illustrates robustness of the definition of inclusion of secondary cases with respect to the association between sample Ct values and transmissibility. Panel a includes secondary cases that tested positive 1-14 days after the primary case and is the same as Figure 1. Panel b includes secondary cases that tested positive 2-14 days after the primary case. Panel c includes secondary cases that tested positive 3-14 days after the primary case. Panel d includes secondary cases that tested positive 4-14 days after the primary case. The number of primary cases, potential secondary cases, and positive secondary cases, included in each panel can be found in Table S5. The shaded area shows the 95% confidence bands clustered on the household level.

**Figure S8:**
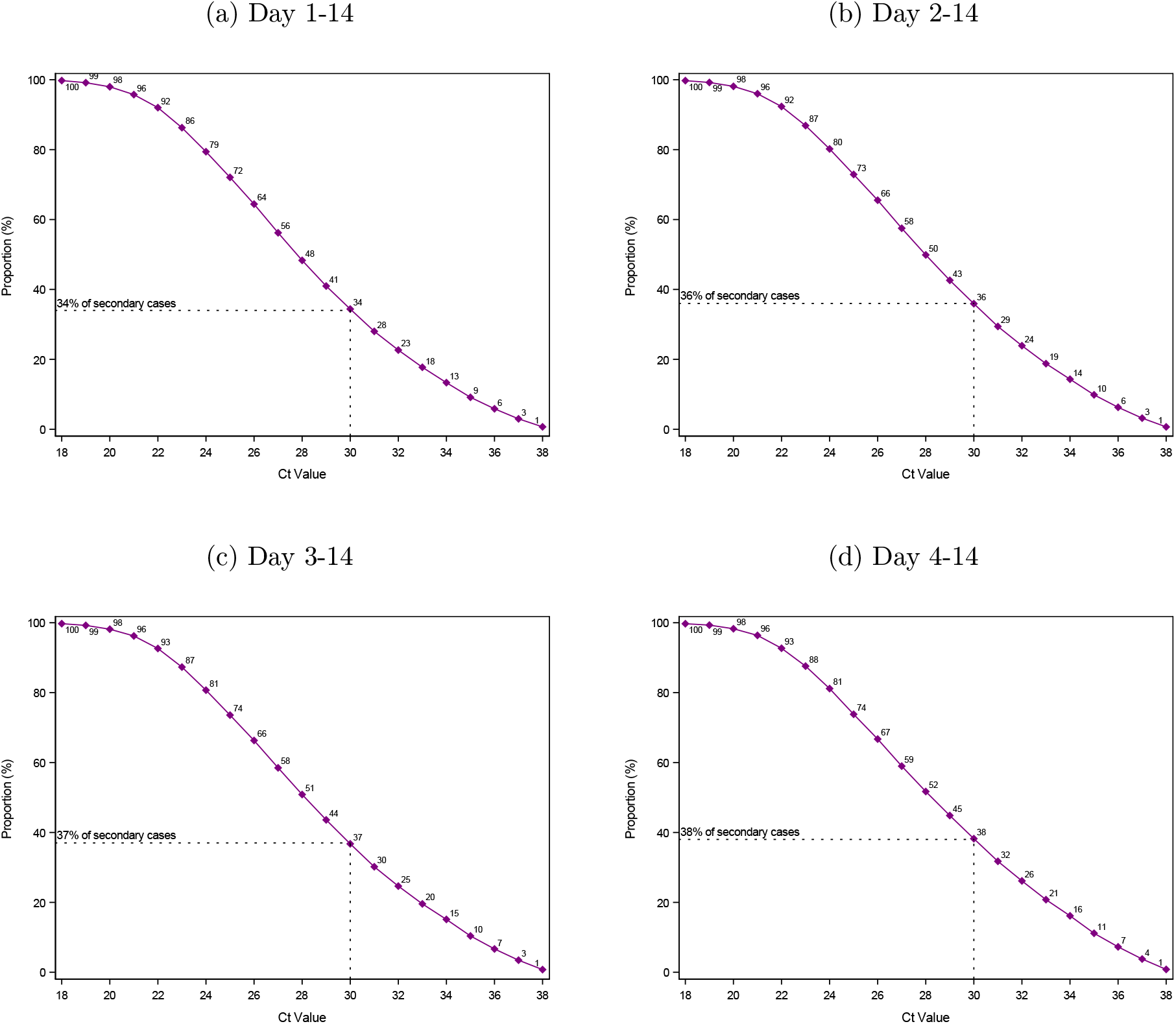
Sensitivity to definition of secondary cases: Proportion of secondary cases associated with Ct values of primary cases above a threshold Notes: This figure illustrates robustness of the definition of inclusion of secondary cases with respect to the reverse cumulative distribution of secondary cases by sample Ct values. Panel a includes secondary cases that tested positive 1-14 days after the primary case and is the same as Figure 2. Panel b includes secondary cases that tested positive 2-14 days after the primary case. Panel c includes secondary cases that tested positive 3-14 days after the primary case. Panel d includes secondary cases that tested positive 4-14 days after the primary case. The number of primary cases, potential secondary cases, and positive secondary cases, included in each panel can be found in Table S5.

**Figure S9:**
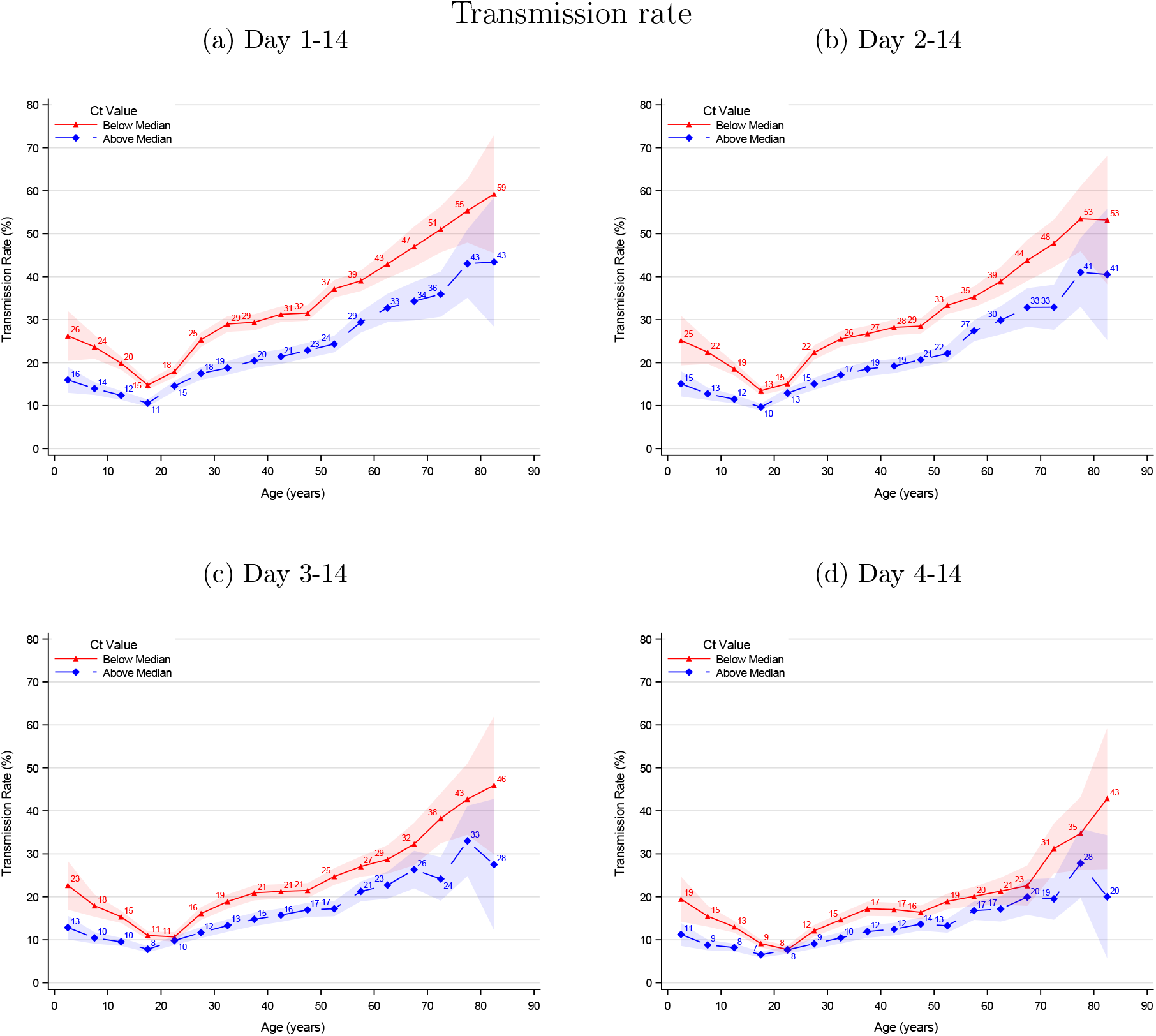
Sensitivity to definition of secondary cases: Age structured transmission rate stratified by median Ct value Notes: This figure illustrates robustness of the definition of inclusion of secondary cases with respect to the age structured transmission rate stratified by median sample Ct value. Panel a includes secondary cases that tested positive 1-14 days after the primary case and is the same as Figure 3, panel a. Panel b includes secondary cases that tested positive 2-14 days after the primary case. Panel c includes secondary cases that tested positive 3-14 days after the primary case. Panel d includes secondary cases that tested positive 4-14 days after the primary case. The number of primary cases, potential secondary cases, and positive secondary cases included in each panel can be found in Table S5. The shaded area shows the 95% confidence bands clustered on the household level.

**Figure S10:**
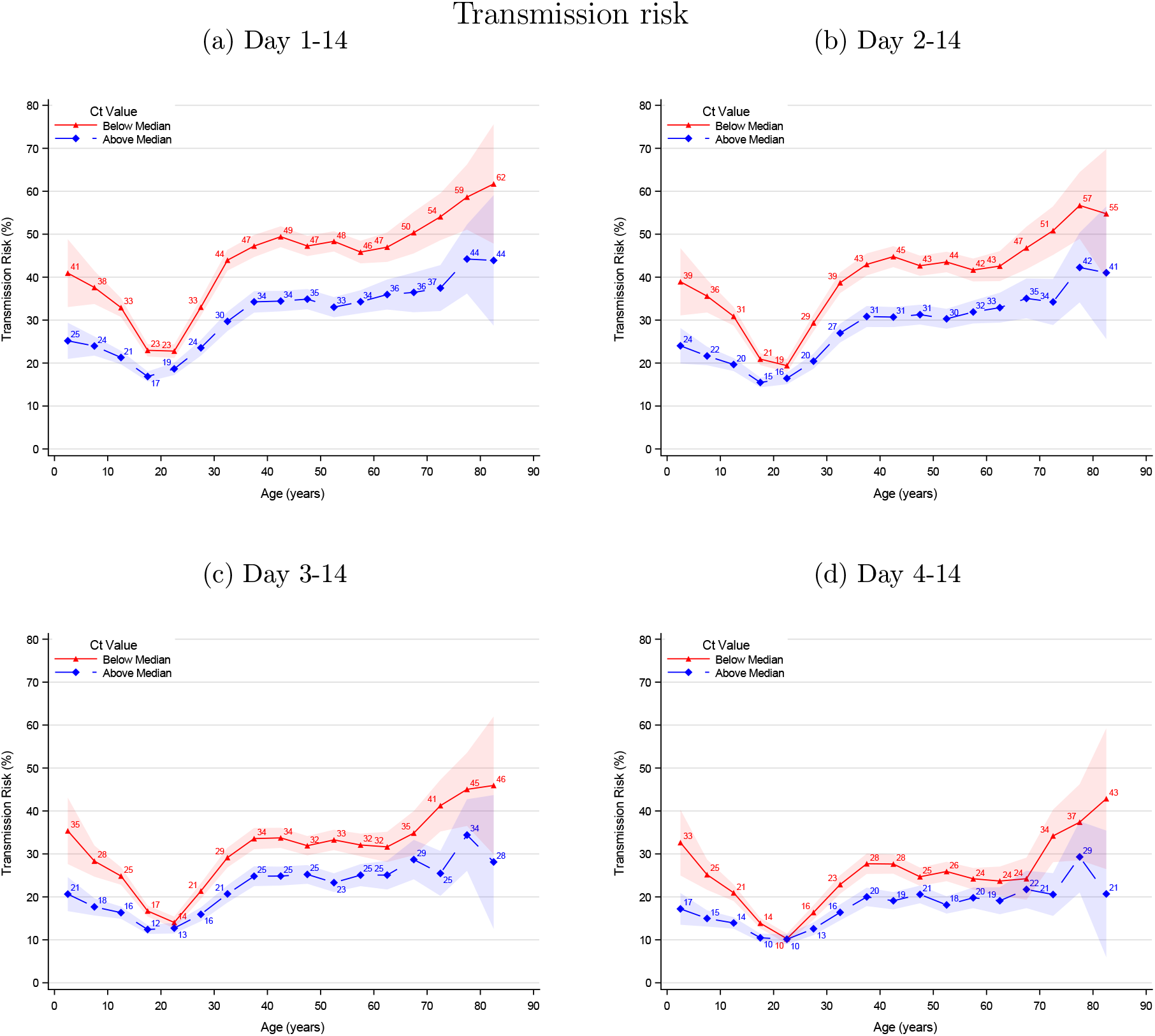
Sensitivity to definition of secondary cases: Age structured transmission rate stratified by median Ct value Notes: This figure illustrates robustness of the definition of inclusion of secondary cases with respect to the age structured transmission risk stratified by median sample Ct value. Panel a includes secondary cases that tested positive 1-14 days after the primary case and is the same as Figure 3, panel b. Panel b includes secondary cases that tested positive 2-14 days after the primary case. Panel c includes secondary cases that tested positive 3-14 days after the primary case. Panel d includes secondary cases that tested positive 4-14 days after the primary case. The number of primary cases, potential secondary cases, and positive secondary cases included in each panel can be found in Table S5. The shaded area shows the 95% confidence bands clustered on the household level.

**Figure S11:**
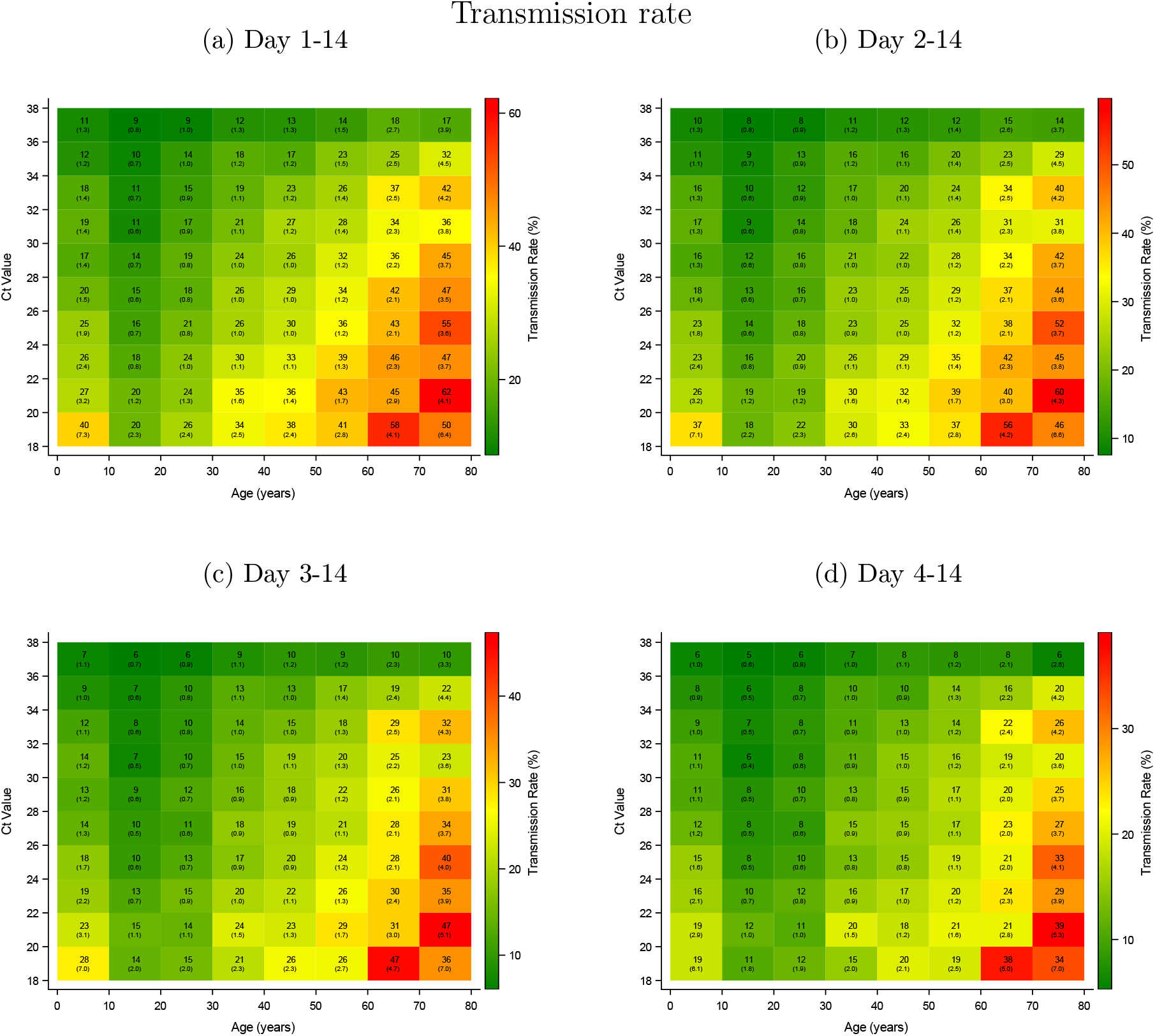
Sensitivity to definition of secondary cases: Association between age, Ct value, and transmission rate Notes: This figure illustrates robustness of the definition of inclusion of secondary cases with respect to the association between age, sample Ct value, and transmission rate. Panel a includes secondary cases that tested positive 1-14 days after the primary case and is the same as Figure 4. Panel b includes secondary cases that tested positive 2-14 days after the primary case. Panel c includes secondary cases that tested positive 3-14 days after the primary case. Panel d includes secondary cases that tested positive 4-14 days after the primary case. The number of primary cases, potential secondary cases, and positive secondary cases included in each panel can be found in Table S5. Standard errors clustered on the household level in parenthesis.

**Figure S12:**
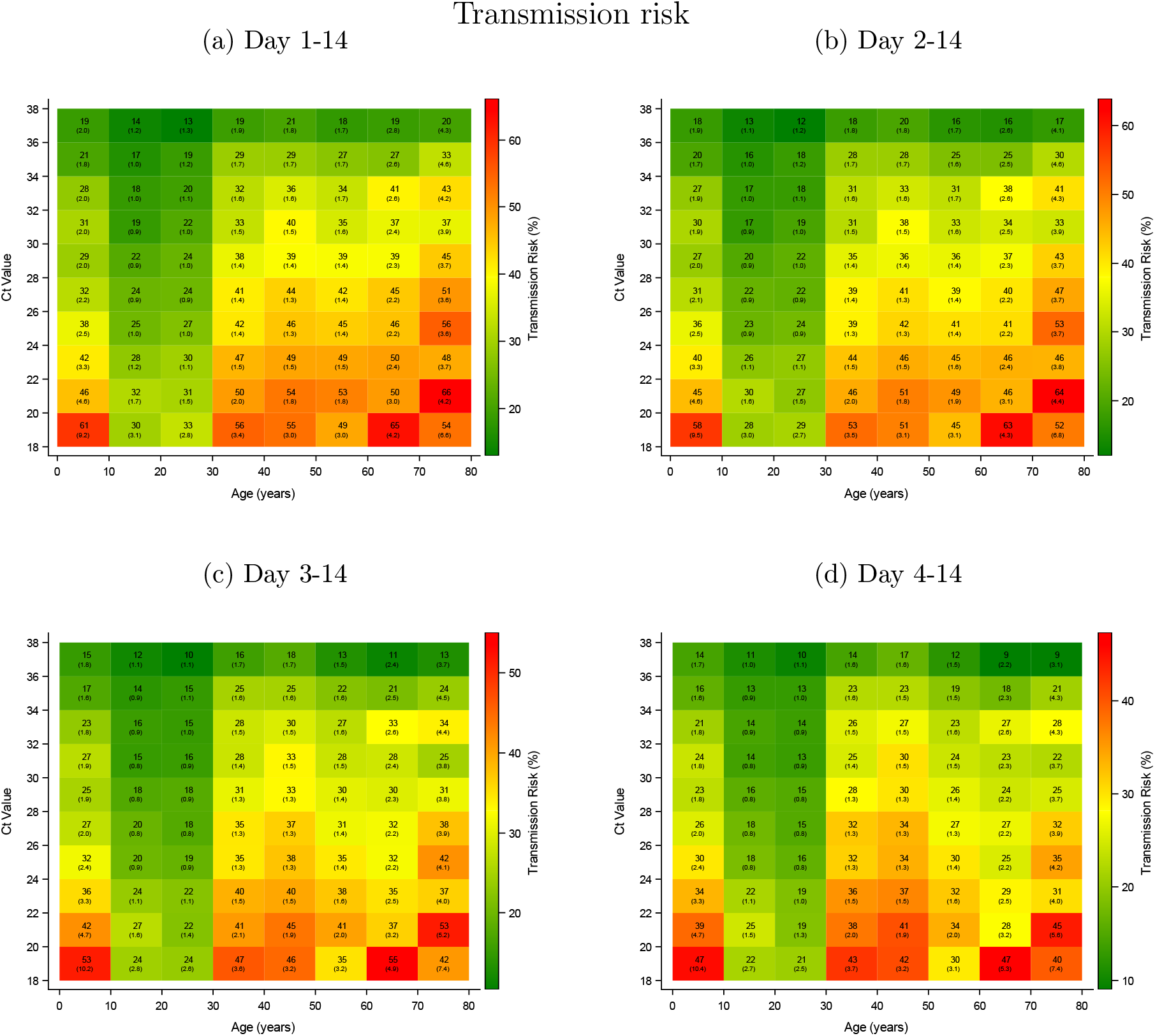
Sensitivity for definition of secondary cases: Association between age, Ct value, and transmission risk Notes: This figure illustrates robustness of the definition of inclusion of secondary cases with respect to the association between age, sample Ct value, and transmission risk. Panel a includes secondary cases that tested positive 1-14 days after the primary case and is the same as Figure 4. Panel b includes secondary cases that tested positive 2-14 days after the primary case. Panel c includes secondary cases that tested positive 3-14 days after the primary case. Panel d includes secondary cases that tested positive 4-14 days after the primary case. The number of primary cases, potential secondary cases, and positive secondary cases included in each panel can be found in Table S5. Standard errors clustered on the household level in parenthesis.

**Figure S13:**
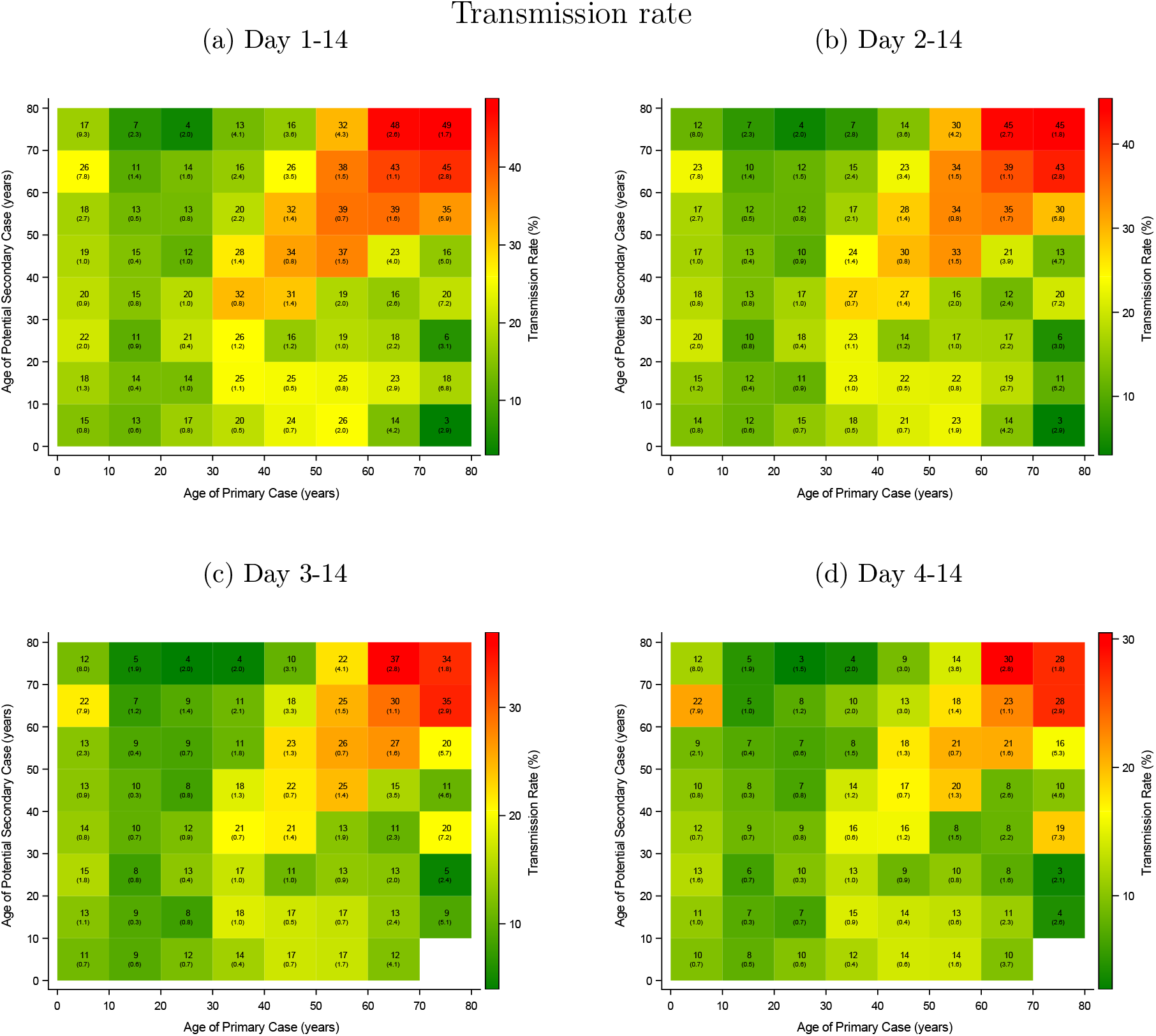
Sensitivity to definition of secondary cases: Association between age of primary case, potential secondary case, and transmission rate Notes: This figure illustrates robustness of the definition of inclusion of secondary cases with respect to the association between age of primary case, potential secondary case, and transmission rate. Panel a includes secondary cases that tested positive 1-14 days after the primary case. Panel b includes secondary cases that tested positive 2-14 days after the primary case. Panel c includes secondary cases that tested positive 3-14 days after the primary case. Panel d includes secondary cases that tested positive 4-14 days after the primary case. The number of primary cases, potential secondary cases, and positive secondary cases included in each panel can be found in Table S5. Standard errors clustered on the household level in parenthesis.

## Appendix D

### Statistical analyses

To estimate the association between Ct values and transmissibility (*y*_*p*_), we estimated the non-parametric regression equation:

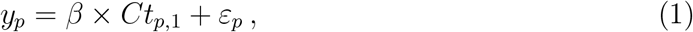

where *Ct*_*p*,1_ is the sample Ct value (rounded to the nearest integer) of the primary case. *β* measures the transmission risk for each Ct value. *ε*_*p*_ denotes the error term, clustered on the household (event) level. The regression was weighted on the primary case level, so each primary case has a weight of one.

To estimate the association between age and transmissibility, stratified by the median Ct value (28), we estimated the non-parametric regression equation:

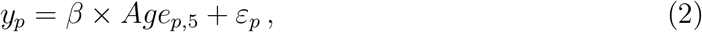

where *Age*_*p*,5_ is the age (in five-year groups) of the primary case. *β* measures the transmissibility for each five-year age group of the primary cases. *ε*_*p*_ denotes the error term, clustered on the household (event) level. The regression was weighted on the primary case level, so each primary case has a weight of one.

To estimate the association between Ct value, age, and transmissibility, we estimated the non-parametric regression equation:

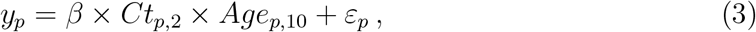

where *Ct*_*p*,2_ is the Ct value (in bi-value groups) and *Age*_*p*,10_ is the age (in ten-year groups) of the primary case. *β* measures the transmission risk of the interaction between Ct value and age of the primary case. *Age*_*s*_ is the age of the potential secondary cases (*s*). *ε*_*p*_ denotes the error term, clustered on the household (event) level. The regression was weighted on the primary case level, so each primary case has a weight of one.

To quantify the increased transmissibility across different observable characteristics, we estimated a univariable and a multivariable logistic regression. In particular, to estimate the odds ratio, we estimated the logistic regression equation with the following linear predictor:

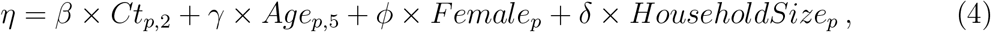

where *β* measures the non-parametric association with Ct values, *γ* measures the non-parametric association with age of the primary case, *φ* measures the association with sex, and *δ* measures the association with the size of the household. *ε*_*p*_ denotes the error term, clustered on the household (event) level. The regression was weighted on the primary case level, so each primary case has a weight of one.

